# Where has democracy helped the poor? Democratic transitions and early-life mortality at the country level

**DOI:** 10.1101/2020.09.09.20191502

**Authors:** Antonio Pedro Ramos, Martiniano Jose Flores, Michael L. Ross

## Abstract

The effects of democracy on living conditions among the poor are disputed. Previous studies have addressed this question by estimating the average effect of democracy on early-life mortality across all countries. We revisit this debate using a research design that distinguishes between the aggregated effects of democracy across all countries and their individual effects within countries. Using Interrupted Time Series methodology and estimating model parameters in a Bayesian framework, we find the average effect of democracy on early-life mortality to be close to zero, but with considerable variation at the country-level. Democratization was followed by fewer child deaths in 21 countries, an increase in deaths in eight, and no measurable changes in the remaining 32 cases. Transitions were usually beneficial in Europe, neutral or beneficial in Africa and Asia, and neutral or harmful in Latin America. The distribution of country-level effects is not consistent with common arguments about the conditional effects of democratic transitions. Our results open a new line of research into the sources of theses heterogeneous effects.

## 1 Introduction

Many studies suggest that democracies are better than autocracies at improving living conditions for the poor. If this is true, countries that transit from autocracy to democracy should see faster reductions in early life mortality (neo-natal, infant, and child mortality) — the most widely-used indicators for the overall well-being of low-income communities^∗^. Previous studies, however, report mixed results: some find that on average, democratic transitions are followed by lower infant and child mortality rates (Przeworski et al., 2000; Lake and Baum, 2001; Gerring et al., 2012a; Wigley and Akkoyunlu-Wigley, 2011; Wang et al., 2018; Krueger et al., 2015), yet others claim that democratic transitions make little or no difference (Mulligan et al., 2004; Ross, 2006).

Our study addresses two important limitations of earlier research. First, most prior studies focus on the average effect of democracy across countries, which tells us little about whether the treatment had significant effects on any of the countries – a question that both scholars and policymakers care deeply about. There is also no reason to assume that the introduction of democratic regimes in different countries will have similar effects on early-life mortality. In fact, many studies explicitly or implicitly suggest the effects of democratization on human welfare are conditional on other factors like state capacity, the income of the median voter, or the availability of international aid.

To capture the heterogeneous effects of democracy at the country level we use a Bayesian hierarchical model that allows us to estimate the effect of each one of the 61 sufficiently long transitions in our data. Our approach compares the observed post-transition mortality rates to counterfactuals constructed from each country’s prior trajectory. We interpret the difference between the observed outcome and the counterfactual as democratization’s net effect.

The second limitation is that past studies have conflated the short and long term effects of democratic transitions. Many transitions are accompanied by social upheaval, economic disruptions, or civil wars that could drive up child mortality rates in the short run. These short-term costs would obscure the long-term benefits of living under democratic rule.

To address this problem, for each of the 61 transitions we decompose changes in mortality after the introduction of democracy into short term shifts in their levels and long term changes in their trends. We use an Interrupted Time Series (ITS) methodology, with model parameters estimated in a Bayesian framework (Fitzmaurice et al., 2011; Singer and Willett, 2003; Morgan and Winship, 2019; Kontopantelis et al., 2015; Turner et al., 2019; Bernal et al., 2017). We then use the estimates of the short and long term effects to construct counterfactual mortality trends, which represent how the mortality rates would have looked in the absence of a democratic transition, and use these trends to estimate how many lives were saved or lost due to democratization. In the main text we present and discuss the effects of democracy on child mortality. In the appendix we show that these results are similar for both neonatal and infant mortality. Our analysis yields three findings.

First, while the average effect of democracy on child mortality across all countries is neither statistically nor substantively significant there is substantial country-level heterogeneity in the effects: democracy was followed by more rapid reductions in child mortality in 21 countries, slower reductions in 8 countries, and no measurable change in the remaining 32 cases. These impacts were sometimes large: for example, we find that in Madagascar, the number of deaths that occurred after democratization was 29% below what it would have been under authoritarian rule. In contrast, we find that in Chile, the number of child deaths increased by almost 50% relative to what we would have expected under authoritarian rule. Most of the large gains in child health were in Sub-Saharan Africa and the post-Communist states of Central and Eastern Europe. Most of the losses were in Latin America, which is surprising given that transitions to democracy in Latin American countries were generally followed by increased spending on social welfare programs.

Second, we find that democratic transitions sometimes have transient short term effects: transitions were helpful for child health in the short-run in four countries, harmful in eight countries, and had no discernible impact in the other 49 cases. Short term harmful effects were concentrated among formerly communist countries, where these transitions were accompanied by economic turmoil and a collapse in social services. These short-run effects, however, were almost always neutralized or reversed by countervailing long-run effects. Short and long term effects are only weakly correlated, which suggests the importance of quantifying each effect separately.

Finally, the distribution of country-level effects is not consistent with common arguments about the conditional effects of democratic transitions due to state capacity, corruption, type of political institutions, income levels, prior democratic history, freedom of the press, foreign aid, or armed conflict. While the patterns are clustered at a regional level, we cannot tell if this is due to country-to-country spillover effects, or shared but unobserved regional characteristics. Indeed, our capacity to make inferences about any conditional effects is limited. We encourage other scholars to study this issue more carefully.

Collectively these findings suggest a new perspective on the welfare effects of democratic transitions. Most prior research has focused on the size and statistical significance of the average treatment effect across all countries. We demonstrate that the average treatment effect tells us little about democratization’s impact at the country level, where the consequences vary widely. Our analysis also finds that transitions have no measurable effect on child mortality trends in about half of all cases, beneficial effects in one-third of the countries and harmful effects in the remaining one-sixth. Democratization is usually neutral or good for the poor - a result that is not apparent when democracy’s effects are pooled across countries.

In the next section we summarize recent studies of democracy and child health and highlight four possible explanations for heterogeneous effects. In Section 3 we describe our measures of both democratic transitions and infant mortality, and in Section 4 we introduce our methodology and compare it with previous approaches. In Section 5 we present the results from our main analyses. We conclude in Section 6 by discussing the significance of our key results and their implications for future research. In the appendix, we show that the results for child mortality presented in the main text also hold true for neo-natal and infant mortality, and demonstrate that the pattern of heterogeneity cannot be easily explained by existing theories.

## 2 Democracy and Human Welfare

Political philosophers from Aristotle to Amartya Sen have argued that democratic governments are better than autocracies at meeting the needs of the disadvantaged. In contemporary debates, these claims are usually articulated through three arguments. The first is that democracies enfranchise low-income voters and legalize civic associations that can advocate on their behalf; together, this gives democratic leaders a stronger incentive to satisfy the preferences of the poor (Pateman, 1976; Putman, 1993). According to the second claim, the free press found in democracies provides political leaders with the information they need to avert famines and other disasters for marginalized peoples, and gives poor communities greater access to critical information about public health (Sen, 1999b; Wigley and Akkoyunlu-Wigley, 2011).

The third argument is that democracies produce more public services than non-democracies, and that these goods typically benefit the poor (McGuire and Olson, 1996; Niskanen, 1997; Lake and Baum, 2001; De Mesquita et al., 2002). The most influential version of this claim was developed by Meltzer and Richard (1981), who conceptualize democracy as an extension of the franchise from a wealthy elite to the rest of the citizenry. They stipulate that policies in a democracy are determined by the preferences of the median voter. Since income is distributed unequally, the income of the median voter lies below the mean income, causing her to support higher taxes that will redistribute income downwards. As a result, democracies should have higher taxes, larger governments, and more public services.

Empirical research has repeatedly shown that the third argument is at least narrowly correct: democracies tend to have bigger governments than non-democracies (Lindert, 2004; Aidt et al., 2006; Aidt and Jensen, 2009; Gouveia and Masia, 1998; Tavares and Wacziarg, 2001; Ghobarah et al., 2004; Persson et al., 2003). Many studies also infer that these higher expenditures are beneficial for the poor, based on cross-national correlations between democratic regimes and improvements in nutrition (Blaydes and Kayser, 2011), access to electricity (Min, 2015), and most importantly, reduced infant and child mortality (Przeworski et al., 2000; Lake and Baum, 2001; Houweling et al., 2005; Besley and Kudamatsu, 2006a; Gerring et al., 2012a).

Some studies argue that the impact of democracy on child mortality is ambiguous. Ross (2006) suggests that most cross-national studies fail to account for unobserved country-specific effects, and shows that including them sharply reduces the size and statistical significance of the democracy-child health correlations; he also points out that at least some results appeared to be biased by non-random missing data. Mulligan et al. (2004) and Shandra et al. (2004) also fail to find significant correlations between regime type and early-life mortality, and both Kohli (2003) and Moore and White (2003) cast doubt on the theoretical arguments linking democracy to poverty reductions.

Debates about the relationship between a country’s regime type and early-life mortality are difficult to resolve due to challenges in identifying the causal effect of democracy on child mortality. This is in part due to non-random assignment: political regimes are not randomly assigned to countries, making it difficult to compare democratizers to non-democratizers. Moreover, it is hard to know if any observed correlations between democratization and human welfare are causal: the foundational work of Lipset (1959), for example, suggests that democratization and improvements in social welfare are jointly produced by an underlying process of “modernization,” which if true suggest any correlation between democracy and living conditions is spurious.

We are aware of only one study that explicitly documented the heterogeneous effects of democracy on child health for a global sample of countries (Pieters et al., 2016). They found that the effects of democracy were larger in countries with higher *ex ante* child mortality. However, they were able to study only around half of the transitions that we study and found no political explanation for heterogeneous effects. We revisit this topic with a new methodology for a larger set of transitions.

### 2.1 Child Mortality, Poverty, and Preventable Deaths

Within countries, child mortality is concentrated among births from the poorest families, and poverty reductions are associated with child mortality reductions (Gwatkin, 2004; Klasen, 2008; Houweling and Kunst, 2009; Pritchard and Keen, 2016; Rasella et al., 2019). Health policies seeking to reduce child mortality often target the poorest births (Black et al., 2003; Bryce et al., 2003; Jones et al., 2003; Victora et al., 2003). For example, Cash Transfer Programs (CTP), currently implemented in many low and middle income countries often improve infant and child health among the poorest families (Bassett, 2008; Banerjee et al., 2010). Many neonatal, infant, and child deaths are related to poverty and preventable with treatments and preventive measures that are available even for the poorest countries (Black et al., 2003; Jones et al., 2003)

Early-life mortality (neonatal, infant, and child) rates are the most carefully-measured and widely-available indicators of living conditions among the poor in low and middle income countries. They correlate with other measures of well-being among the poor, such as morbidity rates and access to medical, sanitation and educational services. However, in comparison to early-life mortality rates, these other measures of well-being do not have the same coverage and are often less comparable across countries (Black et al., 2008).

### 2.2 Why could the effect of democracy vary across countries?

Several strands of political science research may offer explanations for the heterogeneous effects of democracy. One strand suggests that democracy‘s impact is conditional on state capacity. For example, Devarajan and Reinikka (2004) argue that higher public spending — whether or not it results from a democratic transition — will only improve health and education outcomes when the government has the capacity to transfer resources to frontline service providers, and to make sure they actually deliver the needed services. Similarly, when autocratic governments are highly corrupt, it will impair the state’s capacity to deliver services to those who are in most need; hence a democratic transition may improve child health, if it also leads to a reduction in corruption.

A second line of research suggests that democracy’s impact depends on the welfare needs of the median voter. Several studies argue that democracy transfers power to the middle class rather than the poor (Aidt and Jensen, 2009; Stigler, 1970; Ross, 2006). Hence if the middle class would benefit from the same type of interventions that help the poor — like access to clean drinking water, basic sanitation, and maternal health care — we should expect more democratic governments to deliver them, leading to corresponding improvements in child health. But if median-income voters already have low child mortality rates and are indifferent to the types of public services that are needed by the poor, democratic transitions might have little or no effect on child mortality rates.

Important studies by Gerring et al. (2012a) and McGuire (2013) alternatively suggest that the beneficial effects of democratic institutions accumulate slowly and might only emerge after many decades. Although we cannot test for long-term effects in our data – most transitions are too recent to reveal these long-term consequences – the argument seems to imply that democratic transitions will be more effective when a country has had prior democratic episodes.

A fourth approach builds on the observation that different types of democratic institutions tend to generate different levels of public services (Persson et al., 2003). For example, electoral systems based on proportional representation are usually associated with larger governments and more redistribution; so are parliamentary governments. Hence it is plausible that democratic transitions would produce more rapid reductions in child mortality if they result in governments elected through proportional representation, or have parliamentary governments rather than presidential ones.

The fifth political factor is freedom of the press, which we note above could yield greater attention to the needs of poor populations (Sen, 1999a; Wigley and Akkoyunlu-Wigley, 2011, 2017). Since press freedom varies among autocracies, this could account for the heterogeneous impact of democratic transitions: when an autocracy with a relatively free press is replaced by a democracy, the smaller gain in press freedom might produce smaller changes in child health.

A sixth possibility is conflict. We know that while some democratic transitions are peaceful, others are a consequence of or followed by political and social turmoil and wars. Thus it is useful to investigate whether conflict-ridden new democracies perform differently than conflict-free ones.

Finally, the effects of transitions could depend on foreign aid programs. Countries that transit to democracy receive on average a substantial increase in foreign aid (Reinsberg, 2015), which could help finance efforts to alleviate poverty. In addition, both Svensson (1999) and Kosack (2003) report that foreign aid is more effective at reducing poverty in democracies than in autocracies.

## 3 Measuring Child Mortality and Democracy

We treat our outcome variable, the transition to democracy, as a discrete event rather than a gradual process, and hence we use the widely-accepted dichotomous measure of democracy that was originally developed by Przeworski et al. (2000) and extended by Cheibub et al. (2010). It covers all countries through 2009 and enables us to focus on the changes in child mortality that follow a democratic transition — that is, the first year in which a country that was previously under autocratic rule is governed by officials who were chosen through contested elections.

Of the 175 countries in our data, 70 countries made at least one transition from autocracy to democracy between 1970 and 2009. We limit our analysis to the 61 countries whose democratic transitions survived for at least five years (Table 1 in the appendix) so that we can more meaningfully estimate the full effect of a democratic transition on under-5 mortality. This limits our sample to countries that transitioned no later than 2004.^†^ For the five countries that made multiple transitions lasting at least five years (Ecuador, Nepal, Pakistan, Paraguay, and Peru) we estimate the effects of the first transition only, reasoning that outcomes during subsequent transitions may be confounded by prior ones.

According to Ross (2006), in prior studies infant and child mortality data have been characterized by nonrandom missingness, and that the estimated relationship between democracy and child health was dependent on the choice of data set. Rajaratnam et al. (2010) described frequent measurement errors and inconsistencies across the most commonly used data sets.

To address this concern we use new early-life mortality data from Rajaratnam et al. (2010) that covers 175 countries from 1970 to 2009 with no missingness and less measurement error than previous data sets. The underlying data are drawn from vital registration systems, summary birth histories, and complete birth histories, and were largely collected by independent international agencies; the data were then combined using Gaussian process regression, which captures the uncertainty caused by sampling and non-sampling error across data types.^‡^

## 4 Identifying Heterogeneous Effects

We use and Interrupted Time Series (ITS) regression model to estimate the effect of the democracy transitions on child mortality and decompose the effect into short and long term effects. We estimate the model parameters in a Bayesian framework to calculate quantities of interest with appropriate uncertainty measures.

### 4.1 Interrupted Time Series Regression

Our modeling strategy starts with an ITS design to evaluate discontinuities in over time trends in early-life mortality after the introduction of democracy in prior non-democratic countries. ITS is a common approach to evaluate the effect of interventions on health outcomes when experimental designs are not possible by making full use of the available longitudinal data to compare pre and post intervention outcomes (Morgan and Winship, 2019; Kontopantelis et al., 2015; Turner et al., 2019; Bernal et al., 2017). An advantage that ITS designs have in comparison to other quasi-experimental designs is that they limit the effects of selection bias and confounding due to between-group differences. This is particularly useful for our study, as we don’t have a clearly defined and stable control group, as the number of treated units (democracies) and untreated units (dictatorships) changes over time. ITS is similar to segmented regression and regression discontinuity designs, and has been widely used in science and health, for example in evaluating the impact of public health policies on COVID-19 (Vokó and Pitter, 2020; Flaxman et al., 2020). Our basic framework for ITS is illustrated in Figure 1, with hypothetical data that mimics actual mortality data on the log-scale.

**Figure 1:**
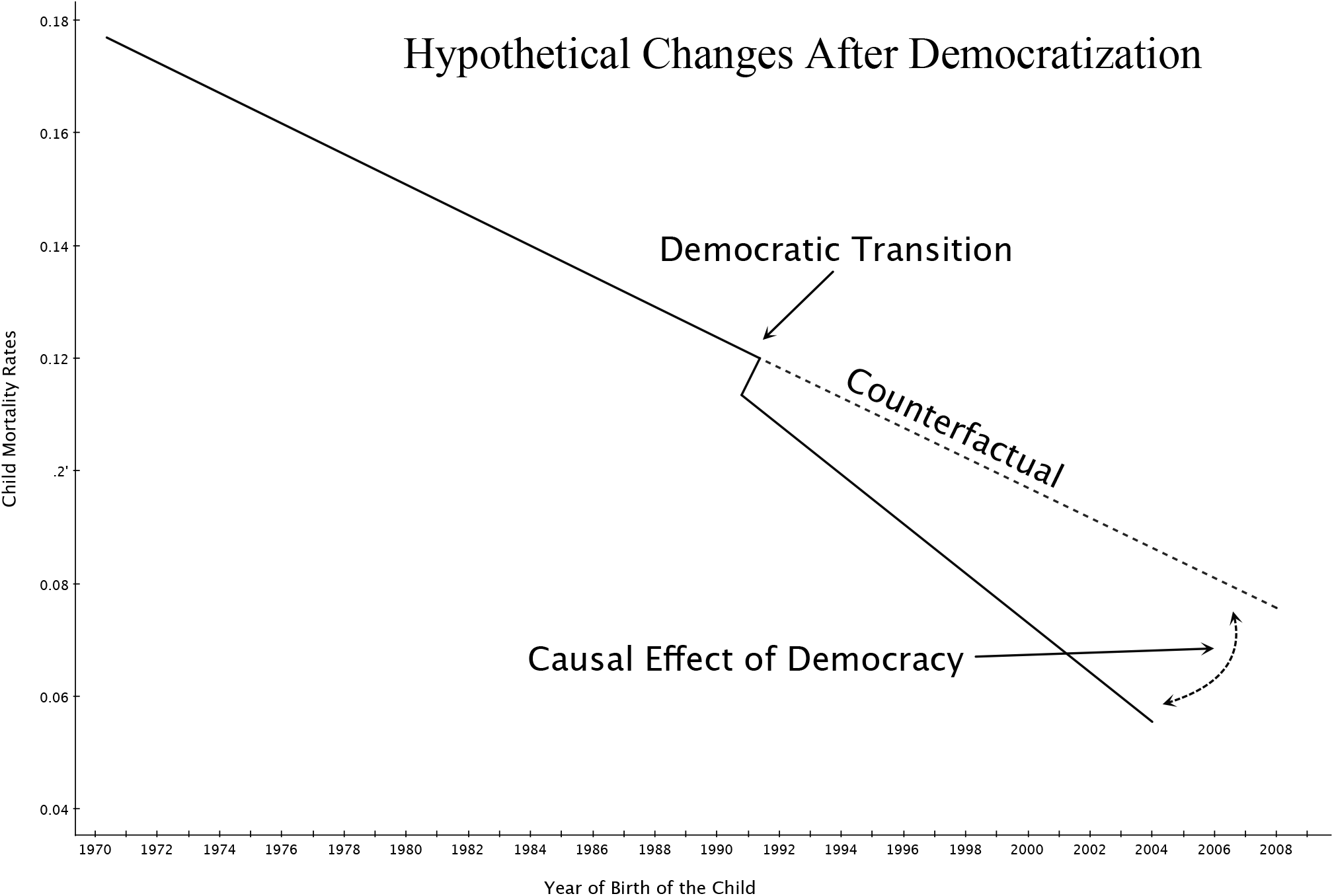
Hypothetical time trends in child mortality for a country that undergoes a single transition from dictatorship to democracy. The dotted line following the transition represents the counterfactual scenario (i.e. without the democratic transition), which is inferred from the pre-transition trend. The effect of democratization is defined as the difference between the observed and counterfactual lines following the year of transition.

The model graphically depicted in Figure 1 can be written as

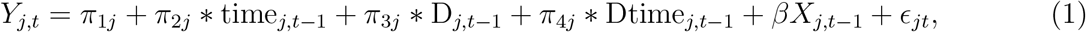

where for *j* = {1,…, 61}, *Y_jt_* is the outcome of interest (neonatal, infant or child mortality on the log scale) for country *j* at time *t*; *X_jt_* are time-varying controls and *β* their coefficients; ∊*_jt_* is the error term. The baseline intercept and the time trends for each country *j* are *π*_1*j*_ and *π*_2*j*_, respectively. To identify the effect of democracy on health, the key variables are *D_jt_*, which is change in the intercept due to the introduction of democracy in the autocratic country, and Dtime_*jt*_ which is a change in slope. Their associated regression coefficients are *π*_3*j*_ and *π*_4*j*_, which represent the magnitudes of the short and long term effects of democracy on child mortality respectively.

Using this model, we can take a country that democratized, and set *D_jt_* and Dtime_*jt*_ to zero to estimate the mortality rate under the counterfactual scenario where the country had never democratized. This allows us to estimate two additional quantities of interest for each transition: (1) the counterfactual trends for child mortality rates in the absence of democracy, and (2) the number of lives saved or lost by the introduction of democracy in the autocratic country.

As with any other statistical methods, ITS relies on some key assumptions. First, the pre and post intervention periods should be clearly defined. This is facilitated by using a dichotomous measure of democracy which is defined based on country’s observable characteristics. Second, the pre-trend period period should be long enough to allow for a good estimation of the post transition trends. In our data, the period of dictatorship is typically longer than the period of democracy, which aids in meeting this assumption. A third assumption is that all time-varying confounders are either absent or controlled for. We discuss this point the next section. Common limitations to ITS approaches include seasonality, which is not an issue with our data, and temporal correlation, which is accounted for by incorporating random effects into the model described below.

### 4.2 Postreatment bias, control variables, and time-varying coun-founders

Since we want to measure the total effect of democracy on early-life mortality, we do not want to control for variables that are themselves affected by the introduction of democracy, because this would introduce post-treatment bias (Gelman and Hill, 2006). For example, previous studies have suggested that democratization also affects GDP per capita, maternal education and HIV prevalence (Acemoglu et al., 2008; Stasavage, 2005; Harding and Stasavage, 2013; Acemoglu et al., 2019; Justesen, 2012). Controlling for these types of variables can potentially lead to an underestimation of the net effect of democracy on early-life mortality. We cannot formally test for post treatment bias or time-varying confounders. Hence our preferred specification for Equation (1) does not include any covariates. However, we show the robustness of our results to the inclusion of these covariates, which suggests that they are not in the causal path between democracy and early-life mortality.

### 4.3 Bayesian random effects models

We use a Bayesian hierarchical model to estimate the effects of democracy on child mortality for each transition simultaneously. Our data are correlated over time within each country. This approach also implements partial pooling, and allows us to generate predictions for the countries’ mortality rates over time (Robinson, 1991; Reisel, 1985; Weiss, 2005; Bell and Jones, 2015). Moreover, since quantities (1) and (2) are functions of the model parameters, point and interval estimates come directly from the posterior samples. We use uninformative priors in our models and thus our results are similar to maximum likelihood estimates for random effects models.

Let *π_j_* = (*π*_1*j*_, π_2*j*_, π_3*j*_, π_4*j*_)^*T*^. We treat *π_j_* as random and estimate it using the following Bayesian hierarchical model,

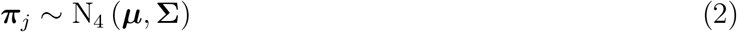

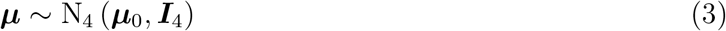

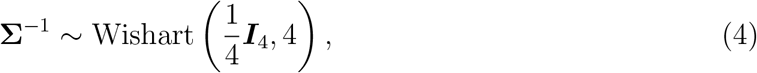

where *μ* is the prior mean vector for *π_j_, I*_4_ is the 4 × 4 identity matrix, Σ is an unstructured covariance matrix, and *μ*_0_ = (−*3*, −0.5, 0, 0) is the mean of the hyperprior for *μ*. The prior distribution for *μ* gives each country a prior mean mortality rate of about 5% at baseline, which decreases multiplicatively by 40% each year. Further, the model says a priori that democracy will have no effect on the mortality rate for any of the countries. However the hierarchical structure allows us to estimate the *π_j_* for each country as well as the correlation between the *π_j_*. Thus, the model will estimate different baseline mortality rates, log-linear trends, and short and long term effects for each country, which also allows us to estimate what the mortality rate would have been had the country never democratized as well as how many lives were saved or lost due to democratization.

## 5 Results

Table 1 presents both the regression coefficients and random effect variances from models that estimate the effect of democracy on child mortality. In all models, credible intervals for the short term and long term fixed effects contain zero, suggesting that the average effect of democracy across all countries is not significant. In column 2 we demonstrate that the inclusion of a standard set of covariates (GDP per capita, maternal education, and HIV prevalence) leaves the short term and long term effects largely unchanged, suggesting that the effects we see are not mediated by these covariates. In the appendix, we also present the main results from the models for neonatal and infant mortality, which are similar to the results for child mortality.

**Table 1:** Results for the fixed and random effects components for four specifications of the hierarchical longitudinal model. Point estimates are presented along with 95 % point-wise credible intervals in parentheses.

| Parameter | No Covariates | Covariates |
| --- | --- | --- |
| Fixed Effects |  |  |
| Intercept | -2.36 (-2.50, -2.22) | -1.52 (-1.68, -1.38) |
| Time | -0.03 (-0.04, -0.03) | -0.03 (-0.03, -0.02) |
| Short Term | 0.01 (-0.026, 0.04) | -0.01 (-0.05, 0.02) |
| Long Term | -0.01 (-0.03, 0.01) | -0.01 (-0.03, 0.01) |
| HIV Prevalence | – | 1.48 (1.32, 1.64) |
| Maternal Education | – | -0.05 (-0.07, -0.04) |
| GDP | – | -0.08 (-0.10, -0.07) |
| Random Effect Variances |  |  |
| Error Variance | 0.005 (0.005, 0.005) | 0.005 (0.005, 0.005) |
| Intercept | 0.85 (0.68, 1.05) | 0.47 (0.37, 0.60) |
| Time | 0.002 (0.001, 0.002) | 0.002 (0.001, 0.002) |
| Short Term | 0.01 (0.01, 0.02) | 0.013 (0.01, 0.02) |
| Long Term | 0.005 (0.004, 0.008) | 0.005 (0.004, 0.008) |

The random effect variances are large in comparison to the average effects, suggesting that there is a substantial amount of heterogeneity between countries. Figure 2 shows the short and long term effects for each country from our random effects specification. Consistent with the results from Table 1, there is a large amount of heterogeneity in the effects of democracy across countries. While the average of the point estimates is centered around zero, we can see that 19% of the countries have statistically significant short term effects and 48% have significant long term effects. Short term effects tend to be larger in magnitude than long term effects since the long term effects are multiplicative in years, and thus accumulate over time. Short and long term effects are not strongly correlated. Table 3 in the appendix explores the results for the short term effect of democracy on child mortality.

**Figure 2:**
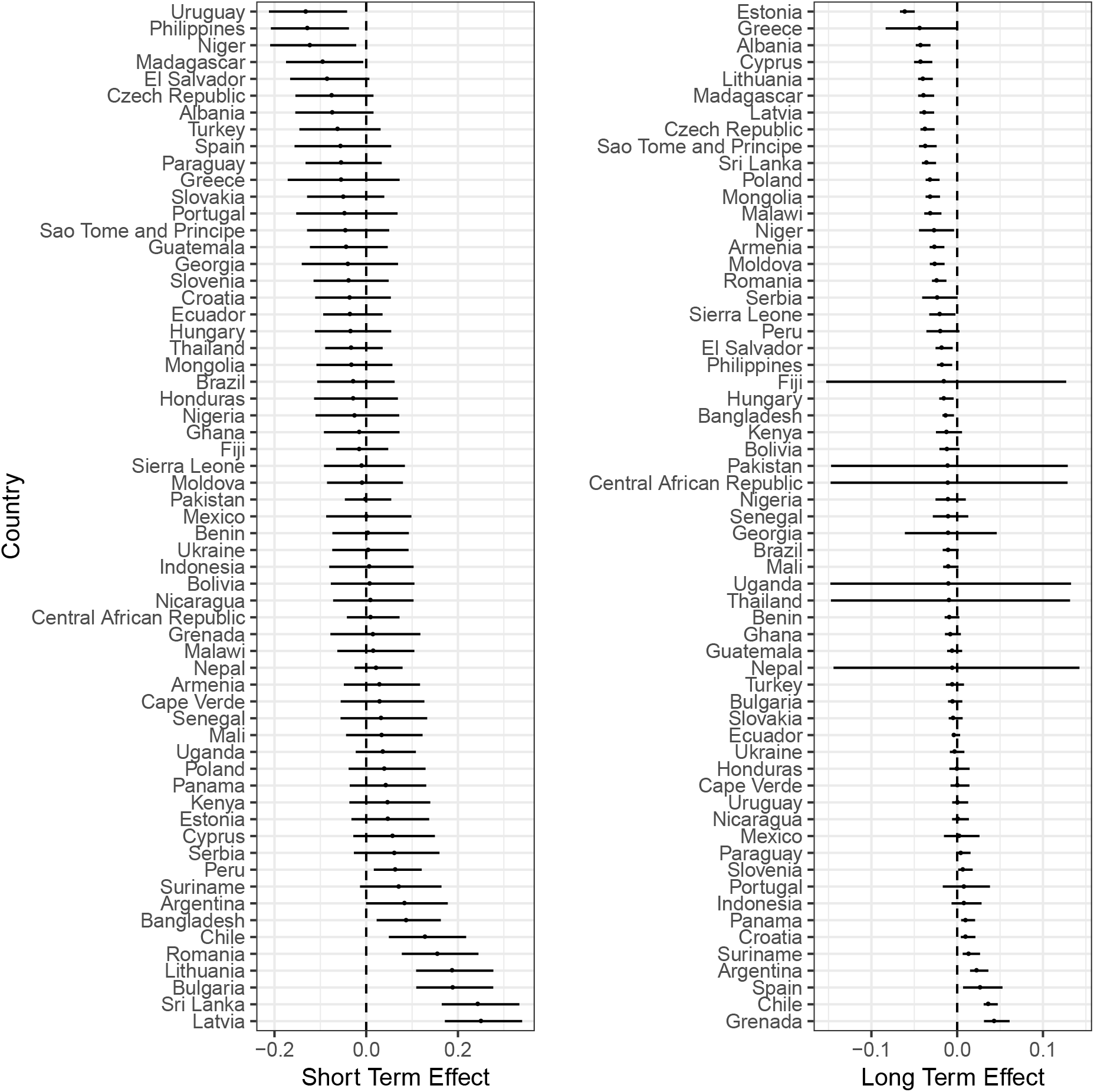
Dot Plots for the short term effects (left) and long term effects (right) from model without covariates, with dotted vertical lines at zero.

In Table 2 we present estimates for the number of lives that were saved or lost after the introduction of democracy for all countries where democratization had statistically significant consequences. We see the largest percentage decrease in the number of lives lost in Estonia and Madagascar, where approximately 1,324 (30%) and 430,000 (30%) of the predicted deaths that would have occurred under authoritarian rule in the respective countries were averted. The largest *total* decrease, measured as the number of deaths averted, was in The Philippines, where an estimated 660,000 lives were saved. Conversely, the largest percentage increase in was in Grenada, which saw an additional 460 child deaths (a 49% increase) under democracy, and the largest *total* increase was in Argentina, where 121,000 additional lives were lost.

**Table 2:** Estimated number of children saved or lost by the introduction of democracy. Results for the model without covariates. Point estimates are presented along with 95 % credible intervals for countries with statistically significant results. Estimates are only for democratic years. Predicted deaths refers to the in-sample predicted number of child deaths under democracy. Counterfactual difference estimates how many more or deaths would have occurred had the country not democratized. Percent difference is the counterfactual difference presented as the proportion of all deaths. Negative numbers for the counterfactual and percent differences mean that democracy was followed by an increase in lives lost.

| Country | Predicted Deaths | Counterfactual Deaths | Lives Saved | Prop |
| --- | --- | --- | --- | --- |
| Grenada | 1,412 (1,370, 1,454) | 953 (796, 1,141) | -458 (-622, -272) | -49.3 (-77.6, -23.8) |
| Chile | 60,570 (58,577, 62,605) | 40,903 (37,293, 44,745) | -19,667 (-23,738, -15,401) | -48.4 (-63.0, -34.7) |
| Argentina | 422,438 (410,652, 434,531) | 301,125 (255,929, 352,731) | -121,314 (-167,385, -69,222) | -41.2 (-65.0, -19.7) |
| Suriname | 7,712 (7,459, 7,975) | 6,277 (5,554, 7,057) | -1,435 (-2,194, -627) | -23.3 (-39.3, -9.0) |
| Bulgaria | 23,967 (23,209, 24,768) | 20,116 (18,210, 22,208) | -3,851 (-5,897, -1,651) | -19.5 (-32.2, -7.4) |
| Panama | 31,597 (30,604, 32,637) | 26,932 (24,165, 30,003) | -4,666 (-7,609, -1,475) | -17.7 (-31.3, -4.9) |
| Peru | 605,057 (578,975, 631,633) | 566,644 (550,316, 583,428) | -38,413 (-70,023, -7,161) | -6.8 (-12.6, -1.2) |
| Sierra Leone | 373,123 (357,208, 389,852) | 408,499 (381,502, 436,402) | 35,375 (4,049, 66,770) | 8.6 (1.1, 15.4) |
| Hungary | 21,334 (20,637, 22,054) | 24,010 (21,768, 26,416) | 2,676 (372, 5,104) | 10.9 (1.7, 19.5) |
| Armenia | 31,132 (30,062, 32,230) | 35,120 (32,045, 38,490) | 3,988 (724, 7,439) | 11.2 (2.3, 19.4) |
| Moldova | 23,662 (22,834, 24,518) | 27,593 (25,229, 30,111) | 3,931 (1,450, 6,528) | 14.1 (5.7, 21.9) |
| Poland | 101,506 (98,102, 104,955) | 119,089 (107,274, 132,149) | 17,583 (5,337, 30,807) | 14.5 (5.0, 23.4) |
| Malawi | 1,120,471 (1,079,936, 1,162,075) | 1,323,452 (1,220,397, 1,435,128) | 202,981 (93,830, 320,821) | 15.2 (7.6, 22.4) |
| El Salvador | 184,121 (178,514, 189,895) | 228,520 (196,800, 265,193) | 44,399 (12,612, 81,222) | 19.0 (6.4, 30.6) |
| Niger | 1,107,476 (1,053,230, 1,164,681) | 1,371,913 (1,288,950, 1,458,836) | 264,437 (167,217, 365,723) | 19.2 (12.8, 25.4) |
| Mongolia | 66,635 (64,354, 68,947) | 83,759 (75,961, 92,212) | 17,123 (9,061, 25,684) | 20.3 (11.9, 28.0) |
| Philippines | 2,093,991 (2,033,754, 2,157,135) | 2,752,621 (2,392,282, 3,167,622) | 658,630 (292,968, 1,074,762) | 23.5 (12.2, 34.0) |
| Cyprus | 2,651 (2,570, 2,734) | 3,562 (3,009, 4,199) | 911 (355, 1,547) | 25.0 (11.8, 37.1) |
| Sao Tome and Principe | 6,876 (6,650, 7,113) | 9,490 (8,340, 10,788) | 2,614 (1,447, 3,935) | 27.2 (17.3, 36.5) |
| Czech Republic | 17,829 (17,238, 18,431) | 24,644 (22,149, 27,343) | 6,815 (4,275, 9,480) | 27.5 (19.3, 34.9) |
| Albania | 26,401 (25,485, 27,354) | 36,807 (33,527, 40,351) | 10,406 (7,048, 13,988) | 28.1 (20.9, 34.9) |
| Madagascar | 1,017,349 (981,159, 1,054,323) | 1,445,821 (1,322,849, 1,577,392) | 428,472 (301,595, 563,900) | 29.5 (22.7, 35.9) |
| Estonia | 3,106 (2,997, 3,218) | 4,430 (4,023, 4,864) | 1,324 (906, 1,771) | 29.7 (22.5, 36.5) |

To help understand how short and long and term effects jointly influence mortality over time, we estimate the predicted mortality rates by year for each country and plot them in Figures 5–8 in the appendix. Those figures compare actual trends with counterfactual trends. We briefly summarize the information from these plots here.

In Central and Eastern Europe, beneficial effects were present in Albania, the Czech Republic, Estonia, Hungary, Moldova, and Poland. In Latvia, Lithuania, Romania, and Bulgaria, the democratic transition led to a long term reduction in the mortality rate over time, but the improvements were partially offset by the short-term costs of the transitions, shown as an immediate spike in the mortality rate. In Bulgaria, the short-term costs were large enough to fully counteract any long-term benefits of democratization.

In five African states (Madagascar, Malawi, Niger, Sierra Leone, and Sao Tome and Principe), democratic transitions were followed by large improvements in child mortality trends. The most dramatic effect was in Madagascar, where the defeat of the ruling socialist party in 1993 led to widespread economic restructuring. In the remaining nine countries, democratization had no discernible impacts. No African state was significantly worse off after its transitions.

In contrast, democratization was less beneficial for child health in Latin America and the Caribbean. Sixteen states in the region became democratic between 1970 and 2009. In five, (Argentina, Chile, Grenada, Panama, Peru, and Suriname), democratization was followed by a deceleration in the reduction of child mortality rates. In the eleven remaining countries, democratic transitions had no statistically significant effects.

Finally, of the remaining sixteen democratizers, transitions were either beneficial or neutral: child mortality rates fell more quickly in five countries (Armenia, Cyprus, Mongolia, The Philippines, and Sri Lanka), and showed no measurable change in the other eleven.

## 6 Explaining Cross-National Results

While our ability to explain the distribution of country-level effects is limited, we analyze the relationships between short and long terms effects with several proxies for the theories discussed on section 2.1. Results are presented on Table 3. The first column lists the variable being analyzed. The second column lists the test type. We use correlations for continuous variables and regression for the categorical variables. Columns 3, 4, 5 and 6 list the effect sizes and associated significance tests. For continuous variables, we present Pearson correlations t-test results. For the categorical variables, we report the *R*^2^ and F-test results. Column 7 lists data sources.

**Table 3:** Democratic History refers to the prior number of democratic years since 1950. Coefficients that are statistically significant at 10 % are in bold.

|  |  | Short | Term | Long | Term |  |
| --- | --- | --- | --- | --- | --- | --- |
| variable | test type | effect | stat.sign | effect | stat.sign | data source |
| Short Term | Correlation |  |  | -0.071 | 0.585 | authors |
| Long Term | Correlation | -0.071 | 0.585 |  |  | authors |
| Corruption | Correlation | -0.231 | **0.081** | 0.197 | 0.139 | V-Dem Project |
| Imp. Adm | Correlation | 0.133 | 0.315 | -0.021 | 0.873 | V-Dem Project |
| GDP per capita | Correlation | -0.018 | 0.889 | 0.011 | 0.932 | V-Dem Project |
| Dem. Hist | Correlation | 0.069 | 0.596 | 0.345 | **0.007** | Bjørnskov & Rode |
| Presidentialism | Correlation | -0.138 | 0.298 | 0.084 | 0.525 | Bjørnskov & Rode |
| Presidential | Regression | 0.017 | 0.319 | 0.016 | 0.337 | V-Dem Project |
| Proportional | Regression | 0.053 | 0.204 | 0.050 | 0.223 | V-Dem Project |
| Prior Regime | Regression | 0.000 | 0.955 | 0.069 | **0.060** | Bjørnskov & Rode |
| Censorship | Correlation | 0.163 | 0.222 | -0.104 | 0.435 | V-Dem Project |
| Battle Intensity | Regression | 0.003 | 0.684 | 0.004 | 0.143 | UCDP |
| Cumulative Battle | Regression | 0.004 | 0.633 | 0.002 | 0.712 | UCDP |

We begin by looking at relationships between the short and long term effects. We find that they are not related, suggesting that our model is useful for disentangling these effects.

Next we turn to the potentially-mediating variables. First, we consider two proxies for state capacity: corruption in the previous regime and impartial administration. We find a marginally-significant correlation between corruption and the short term effects of democratization (p = 0.08), but no long-term effect; there is also no correlation with our measure of impartial administration.

To evaluate the median voter hypothesis, we use a country’s GDP per capita (in the year of transition) but find no correlation with either the short or long term effects of democratization.

We then consider whether the number of prior democratic years – since 1950 but prior to the democratic transition under study – affects the new democratic regime. Having more years under democratic regimes was uncorrelated with the short term effects, but positively correlated with the long-term effects. This implies that when countries have more prior democratic experience – as many in Latin America did – the final democratic transition tended to decrease mortality more slowly over time. This is not consistent with the arguments made by Gerring et al. (2012a) and McGuire (2013) about the long-term effects of democratic experience, but it does account for our findings for the Latin American countries.

We also find no effect from political institutions: there is no difference between countries that transited to presidential or parliamentary systems, or between countries that elected governments through proportional representation or majoritarian systems.

Similarly, there is no relationship between the amount of censorship in the prior regime and short or long term effects, suggesting that freedom of the press is not an important mediator.

To evaluate the impact of post-transition armed conflicts, we consider a five-year period after the transition and distinguish between minor conflicts (between 25 and 1000 battle-related deaths in a given year) and major ones (greater than 1000 battle-related deaths). We also evaluate whether conflicts in the five-year pre-transition period were significant. In each case, we look at both the absolute number of conflicts, and the cumulative number of battle deaths. None of these measures were associated with either short or long term differences in the impact of democratic transitions.

Finally, we consider the effects of both the level of, and changes in, foreign aid. Unlike the other claims, this has a time-series dimension: instead of looking at foreign aid at the moment of transition, we wish to know whether cross-national variations in foreign aid levels, and foreign aid changes, are associated with subsequent variations in child mortality rates. Due to space limitations, results are presented in the appendix. We use the same models used for the main results but add several types of foreign aid as covariates. None of our foreign aid measures are associated with post-transition variations in child mortality rates. See Appendix for full results.

## 7 Discussion

Our study makes two contributions to the long-standing debate over the effects of democratic transitions on the livelihoods of the poor (e.g., Lake and Baum (2001); Ross (2006); Kudamatsu (2012); Besley and Kudamatsu (2006b); Gerring et al. (2012a); Wigley and Akkoyunlu-Wigley (2011)). First, while most previous studies argued about the average effect of democratic transitions, our analysis suggests that this is a relatively uninformative approach. Pooling the effect of democracy across all countries conceals wide variation in country-level effects. Even though our model shows the average effect to be close to zero, we also find that the transition to democracy had a significant effect in almost half of the countries. In those countries where it mattered, democracy produced faster declines in early life mortality almost three-quarters of the time.

These results cast new light on democracy’s regional effects. In most of Central and Eastern Europe, democratic transitions in the 1990s went hand-in-hand with transitions from command to market economies, often-brutal economic shocks, and a rise in poverty, adult disease, and mortality (Unicef, 1997; Safaei, 2012; Bollyky et al., 2019). Our finding that child health also suffered—particularly in Bulgaria, Romania, and the Baltic states—is consistent with this shock. But we also find that once this turbulence passed, child mortality rates fell more quickly than in the final period of Communist rule, suggesting that democratization was good for the poor in about half of these states and had no measurable effect in all but one of the others.

Our Africa results are broadly consistent with earlier studies on the beneficial health effects of Africa’s democratic transitions (Kudamatsu, 2012; Peyvand and Gauri, 2002), but offer a somewhat more complex picture—with better health outcomes for children in one-third of the new democracies and no detectable changes in the remaining two-thirds. Since Africa has the world’s highest child mortality rates, the number of early life deaths averted is large, especially in Madagascar, Malawi, and Niger.

For Latin America, the results are surprising. Earlier studies reported that democratization in Latin America had a strong, positive association with social and health spending (Brown and Hunter, 1999; Huber et al., 2008; Huber and Stephens, 2012; Avelino et al., 2004). Our analysis finds that this additional spending did not improve health outcomes, and that in five cases – Argentina, Peru, Chile, Panama, and Grenada – democratization was followed by a slowdown in the previous rate of decline in child mortality rates. These results echo the cross-national finding of (Filmer and Pritchett, 1999) that higher public health spending is not significantly associated with better health outcomes.

Our Latin America findings also contrast those of McGuire (2013), which argues that democratic transitions in Argentina, Brazil, and Chile led to reduced child mortality. Yet according to our analysis, Brazil’s 1985 transition was followed by a small but statistically insignificant improvement in child health, while transitions from military rule in Argentina (1983) and Chile (1990) were followed by relatively large and statistically significant slow downs in the child mortality reductions that were in place prior to democratization. This may not have been due to any harm inflicted by Chile or Argentina’s newly-democratic governments, but to a reversion to more typical child health trends following a period of exceptionally rapid improvements during the military dictatorships that preceded them.

Finally, we explore several mechanisms that could mediate these heterogeneous effects. Since the number of transitions is small (61), we do not have a lot of statistical power to identify significant results. We consider it notable, however, that there were no strong patterns that validated earlier claims about the conditional effects of democracy. The only association that met the standard threshold for statistical significance – the correlation between longer previous spells of democratic rule pre-transition, and slower reductions in child mortality over the long-term – are not consistent with prior theories about the benefits of democratization (Gerring et al., 2012b). This is a vital topic for future research.

In the subsection 2.3 of our online appendix, *Comparing Actual and Counterfactual Time Trends*, we provide more context for the effect of democracy on child mortality. We predict mortality rates by country and year and plot these predictions in Appendix Figures 2 – 6. In each figure, we plot the empirical mortality rate (dots) and the counterfactual rate with confidence bands (solid line with shaded region). Plots are arranged by geographic region. Estimates are from equations 1–5 from the paper.

In addition to illustrating the heterogeneity of the effects, these plots show that our model produces good in-sample fit. This is particularly useful for causal interpretation of our results as one of the assumptions of the ITS is a good in-sample fit for the pre-democratic period. This allows for a more plausible causal interpretation of our counterfactual extrapolation.(Morgan and Winship, 2019; Kontopantelis et al., 2015; Turner et al., 2019; Bernal et al., 2017).

## 8 Conclusion

Our study shares two important limitations with other cross-national studies on regime type and health. First, our reliance on observational data makes it difficult to make strong claims about causal relationships: we cannot rule out endogeneity, nonrandom selection, and unobserved confounders. And second, our results are sensitive to the assumption that pretransition trends would not have changed in the absence of the democratic transitions. While we cannot know how closely the estimated counterfactual trend matches the “actual” counter-factual trend, our model allows us to quantify our uncertainty, and seems more reasonable than the usual assumption of parallel trends over time. As we show in the appendix, our model also has a very good in-sample fit, suggesting that despite its relatively simple log-linear function fit the data very well.

One benefit of our approach is its relative simplicity. While the random effects structure of our models is complex, the key quantities of interest are estimated using a straightforward functional form with log linear pre-transition timeblue trends and log-linear deviations in the trends after democratization. Similarly, the short term effects are simply intercept shifts after the transition, and long term effects are the post-transition slopes. We refrain from using complex functional forms such as splines that could potentially lead to overfitting; we also exclude possible confounders from our preferred models to avoid any post-treatment biases. Finally the Bayesian approach used in this paper allows us to use well-established statistical theory to calculate point and uncertainty estimates of counterfactual scenarios for countries’ mortality rates under different regimes types and how many lives were saved or lost due to the democratic transitions.

Our findings support a growing literature on the heterogeneous effects of democracy on a variety of outcomes (Olper et al., 2013; de Kadt and Wittels, 2016; Pieters et al., 2016). The fact that these transitions have varied effects should not be a surprise: these transitions have occurred under a wide range of conditions, in different regions and in different periods, and led to the installation of different types of democratic institutions. We should not assume that democratization has similar effects in every setting. Understanding the reasons for these cross-country differences will be an important topic for future research.

## Data Availability

The data is public available

## Acknowledgments

We would like to thank Barbara Geddes, Jeffrey Lewis, Mark Handcock, Chad Hazlett, Robert Weiss and Patrick Heuveline for commenting on previous versions of this paper. We acknowledge financial support from the Eunice Kennedy Shriver National Institute Of Child Health & Human Development of the National Institutes of Health under Award Number K99HD088727.

## Online Appendices for

### 1 Detailed Data Summaries

We have data on 145 independent countries going back to 1970. Of these, 30 (21%) started as stable democracies, 108 (74%) started and stayed as dictatorships, and 7 (5%) started as unstable democracies. Over the next 38 years, 30 new independent governments formed (Table 6). Of these, 12 (40%) started and stayed as democracies, 16 (53%) started as dictatorships, and 2 (6%) started as democracies and transited back to dictatorships. In total, we have 124 dictatorships, 47 (38%) of which eventually transited to stable democracies, and 9 unstable democracies, 8 (89%) of which went on to become stable democracies.

Half of the 124 countries that started as dictatorships in 1970 transited at least once. Of these, 48 transited exactly once, 12 transited twice, one (Suriname) transited three times, and one (Thailand) transited four times. Of the 48 that transited only one time, 39 (41%) of those transitions were into stable democracies. Of the 14 that transited at least twice, 8 (57%) eventually transited into stable democracies. Thus, in total, out of all the countries that underwent at least one period of democracy during the study, 75% became stable democracies indicating that if a country transits at all, it is likely to become to end up as a stable democracy.

For some countries, the health and political data do not match. Countries from the former Soviet Union, such as Ukraine, are counted as separate countries for the child mortality data but as a single entity for the political data. For these cases, we used each country’s individual health data, but kept the regime type the same across all countries.

Yugoslavia is more complex. Separate health information is available for Serbia, Bosnia and Herzegovina, Montenegro, Croatia, and Slovenia, but not for Kosovo. We kept the health data separate, but used the political indicators of Yugoslavia for all countries except for Bosnia and Herzegovina and Montenegro. After the end of the communist rule, we use the coding from Cheibub et al. (2010). We keep information from Montenegro after 2006 and Bosnia and Herzegovina after 1991.

Germany has no separate health data for West and East Germany before the re-unification, yet it is not reasonable to treat both countries as if they were under the same political regime before that time. However, most of the health information was drawn from West Germany, so we also used the political information West Germany before the re-unification. An alternative would be to remove all data from Germany before re-unification but this seems worse than our solution.

Some countries, especially in Africa, were colonies until very recently. Thus, they are not present in these data based on political indicators until some time after 1970, and we included all country-years just after independence from colonial rule. A full list of these countries with their years of independence can be found in table 1. For Vietnam, we have data from the end of the war in 1976.

**Figure 1:**
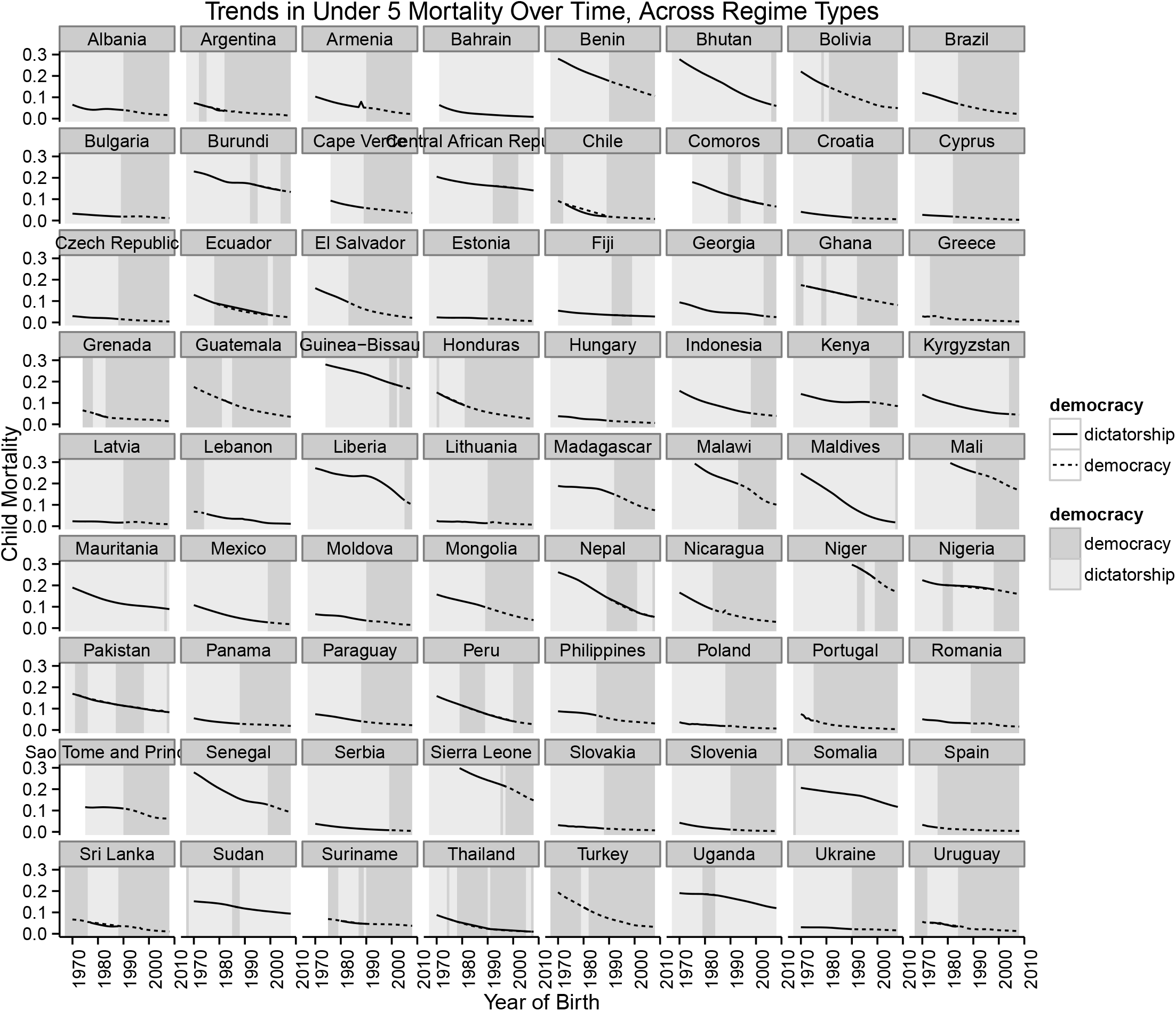
*Trends in Child Mortality for transitional countries using orginal code of democracy from Przeworski at al*.

**Table 1:**
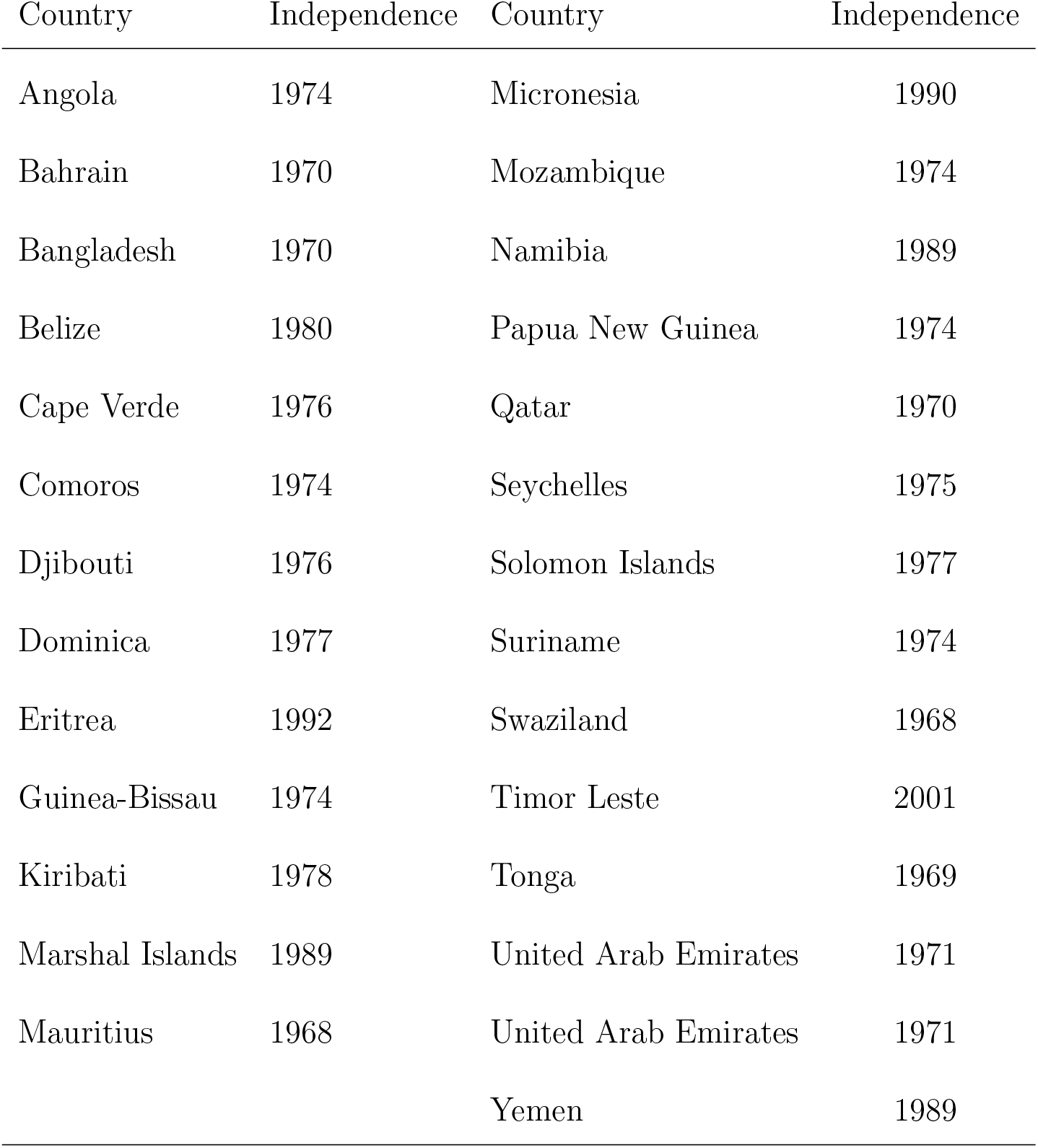
Countries that were not independent in 1970. Source: Cheibub at all (2010)

| Country | Independence | Country | Independence |
| --- | --- | --- | --- |
| Angola | 1974 | Micronesia | 1990 |
| Bahrain | 1970 | Mozambique | 1974 |
| Bangladesh | 1970 | Namibia | 1989 |
| Belize | 1980 | Papua New Guinea | 1974 |
| Cape Verde | 1976 | Qatar | 1970 |
| Comoros | 1974 | Seychelles | 1975 |
| Djibouti | 1976 | Solomon Islands | 1977 |
| Dominica | 1977 | Suriname | 1974 |
| Eritrea | 1992 | Swaziland | 1968 |
| Guinea-Bissau | 1974 | Timor Leste | 2001 |
| Kiribati | 1978 | Tonga | 1969 |
| Marshal Islands | 1989 | United Arab Emirates | 1971 |
| Mauritius | 1968 | United Arab Emirates | 1971 |
|  |  | Yemen | 1989 |

**Table 2:** List of countries with year of stable democratic transition.

| Country | Transition Year | Country | Transition Year |
| --- | --- | --- | --- |
| Albania | 1991 | Mali | 1992 |
| Argentina | 1983 | Mexico | 2000 |
| Armenia | 1991 | Moldova | 1991 |
| Bangladesh | 1986 | Mongolia | 1990 |
| Benin | 1991 | Nepal | 1990 |
| Bolivia | 1982 | Nicaragua | 1984 |
| Brazil | 1985 | Niger | 2000 |
| Bulgaria | 1990 | Nigeria | 1999 |
| Cape Verde | 1990 | Pakistan | 1988 |
| Central African Republic | 1993 | Panama | 1989 |
| Chile | 1990 | Paraguay | 1989 |
| Croatia | 1991 | Peru | 1980 |
| Cyprus | 1983 | Philippines | 1986 |
| Czech Republic | 1989 | Poland | 1989 |
| Ecuador | 1979 | Portugal | 1976 |
| El Salvador | 1984 | Romania | 1990 |
| Estonia | 1991 | Sao Tome and Principe | 1991 |
| Fiji | 1992 | Senegal | 2000 |
| Georgia | 2004 | Serbia | 2000 |
| Ghana | 1993 | Sierra Leone | 1998 |
| Greece | 1974 | Slovakia | 1989 |
| Grenada | 1984 | Slovenia | 1991 |
| Guatemala | 1986 | Spain | 1977 |
| Honduras | 1982 | Sri Lanka | 1989 |
| Hungary | 1990 | Suriname | 1991 |
| Indonesia | 1999 | Thailand | 1992 |
| Kenya | 1998 | Turkey | 1983 |
| Latvia | 1991 | Uganda | 1980 |
| Lithuania | 1991 | Ukraine | 1991 |
| Madagascar | 1993 | Uruguay | 1985 |
| Malawi | 1994 |  |  |

### 2 Closer Look at Heterogeneous Effects of Democratization on Child Mortality

To help understand how short and long and term effects jointly effect mortality over time, we estimate the predicted mortality rates by year for each country and plot them in Figures 2 – 6. In each figure, we plot the empirical mortality rate (dots) and the counterfactual rate with confidence bands (solid line with shaded region). Beneficial effects are indicated by red counterfactual regions, harmful effects by blue, and neutral effects in grey. Estimates are from equations 1–5 from the paper. The out-of-sample predictions were obtained by setting *D_jt_* and Dtime_*jt*_ from equation 1 to zero.

**Figure 2:**
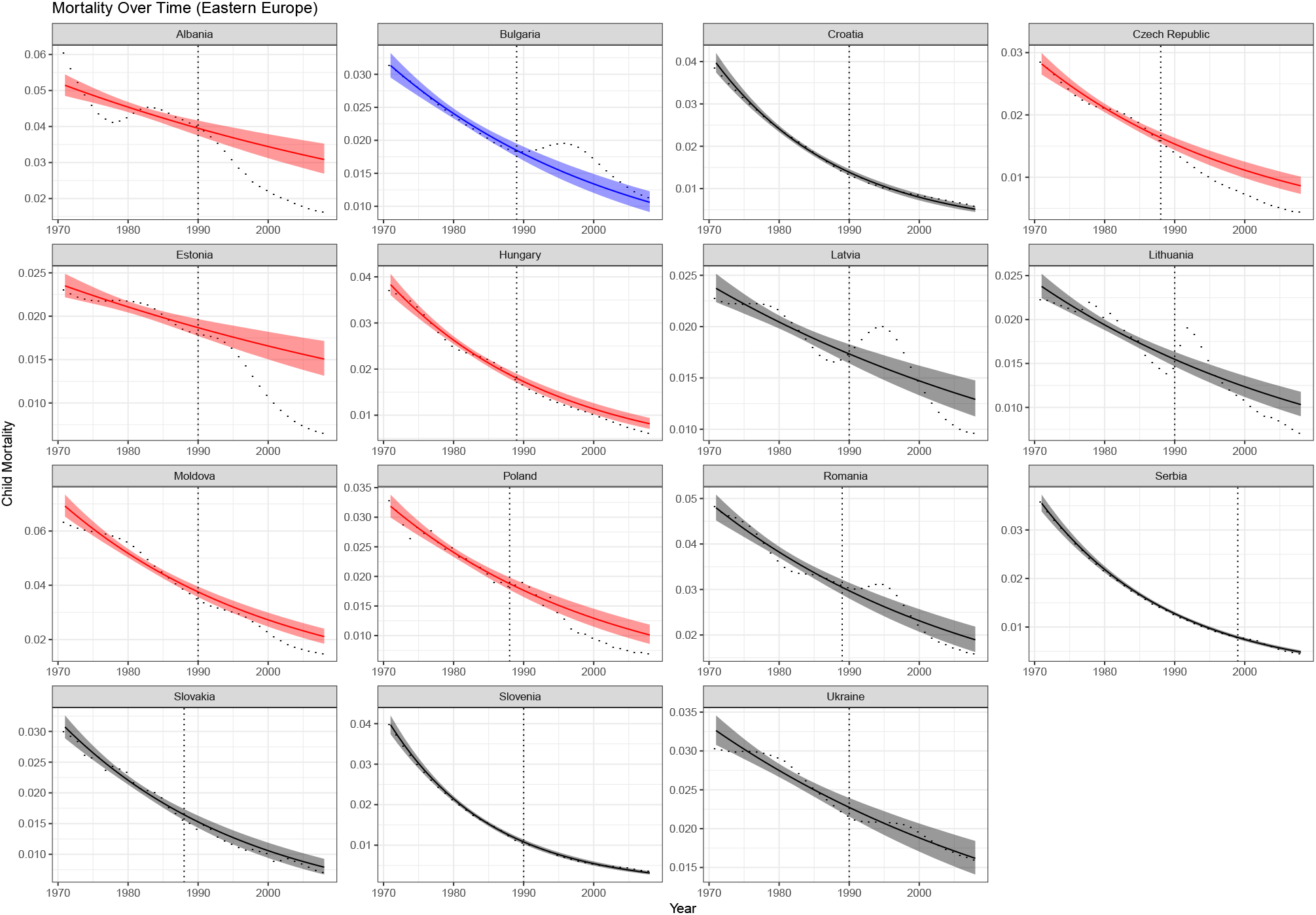
Empirical mortality rate (dots) and counterfactual mortality rates (solid line with shaded region) for European countries. Beneficial effects are indicated by red counterfactual regions, harmful effects by blue, and neutral effects by grey. Dashed vertical line at the year of transition into democracy.

Figure 2 shows the plots for all transitions in Central and Eastern Europe. Beneficial effects were present in Albania, the Czech Republic, Estonia, Hungary, Moldova, and Poland. In Latvia, Lithuania, Romania, and Bulgaria, the democratic transition led to a long term reduction in the mortality rate over time, but the improvements were partially offset by the short-term costs of the transitions, shown as an immediate spike in the mortality rate. In Bulgaria, the short-term costs were large enough to fully counteract any long-term benefits of democratization.

Figure 3 shows the plots for the fourteen African democratizers, where the transitions generally had neutral or beneficial effects. In five African states (Madagascar, Malawi, Niger, Sierra Leone, and Sao Tome and Principe), democratic transitions were followed by large improvements in child mortality trends. The most dramatic effect was in Madagascar, where the defeat of the ruling socialist party in 1993 led to widespread economic restructuring. In the remaining nine countries, democratization had no discernible impacts. No African state was significantly worse off after its transitions.

**Figure 3:**
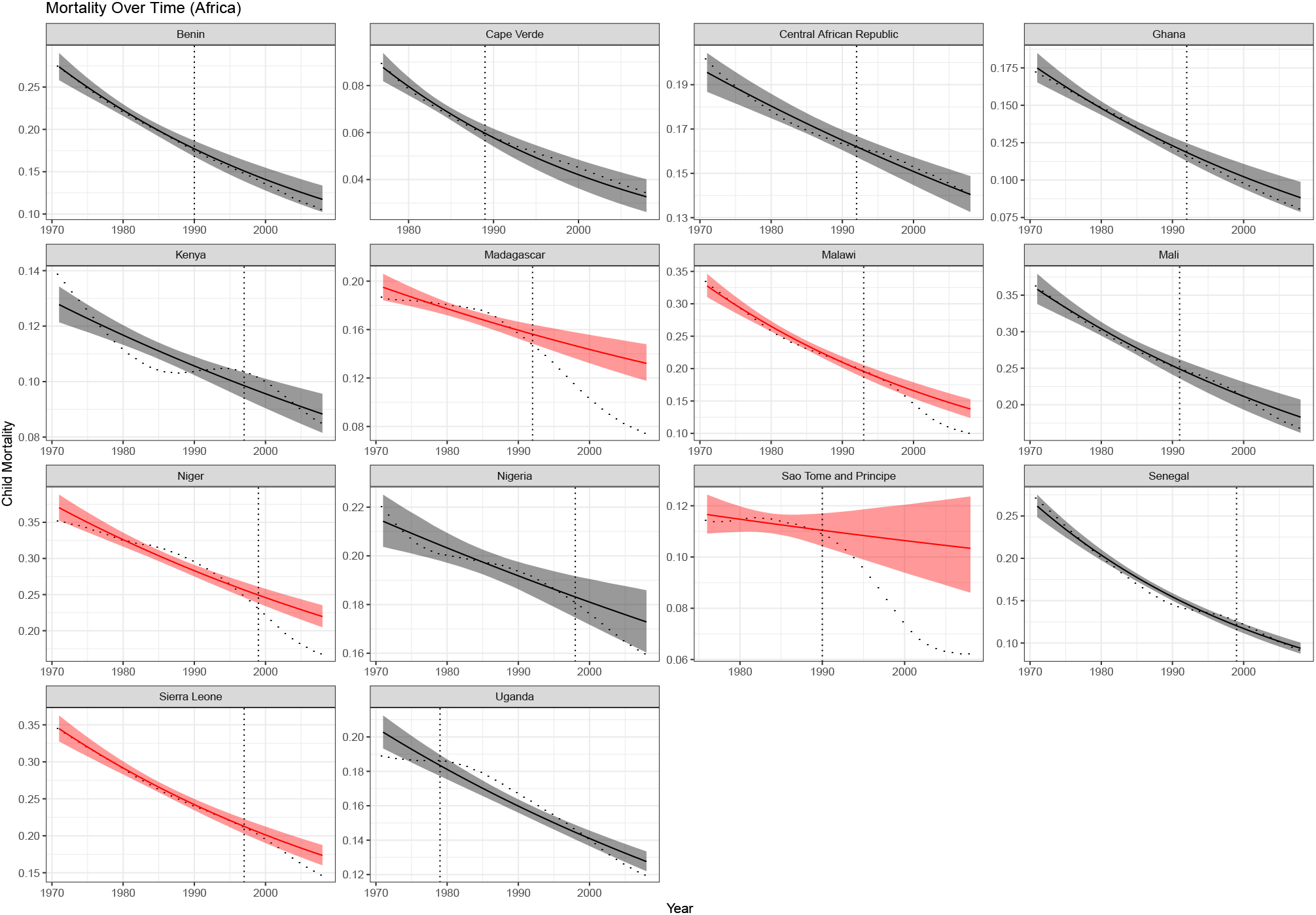
Empirical mortality rate (dots) and counterfactual mortality rates (solid line with shaded region) for African countries. Beneficial effects are indicated by red counterfactual regions, harmful effects by blue, and neutral effects by grey. Dashed vertical line at the year of transition into democracy.

In contrast, democratization was less beneficial for poor communities in Latin America and the Caribbean, as shown in Figure 4. Sixteen states in the region became democratic between 1970 and 2009. In five, (Argentina, Chile, Grenada, Panama, Peru, and Suriname), democratization was followed by a deceleration in the reduction of child mortality rates. In the eleven remaining countries, democratic transitions had no statistically significant effects.

**Figure 4:**
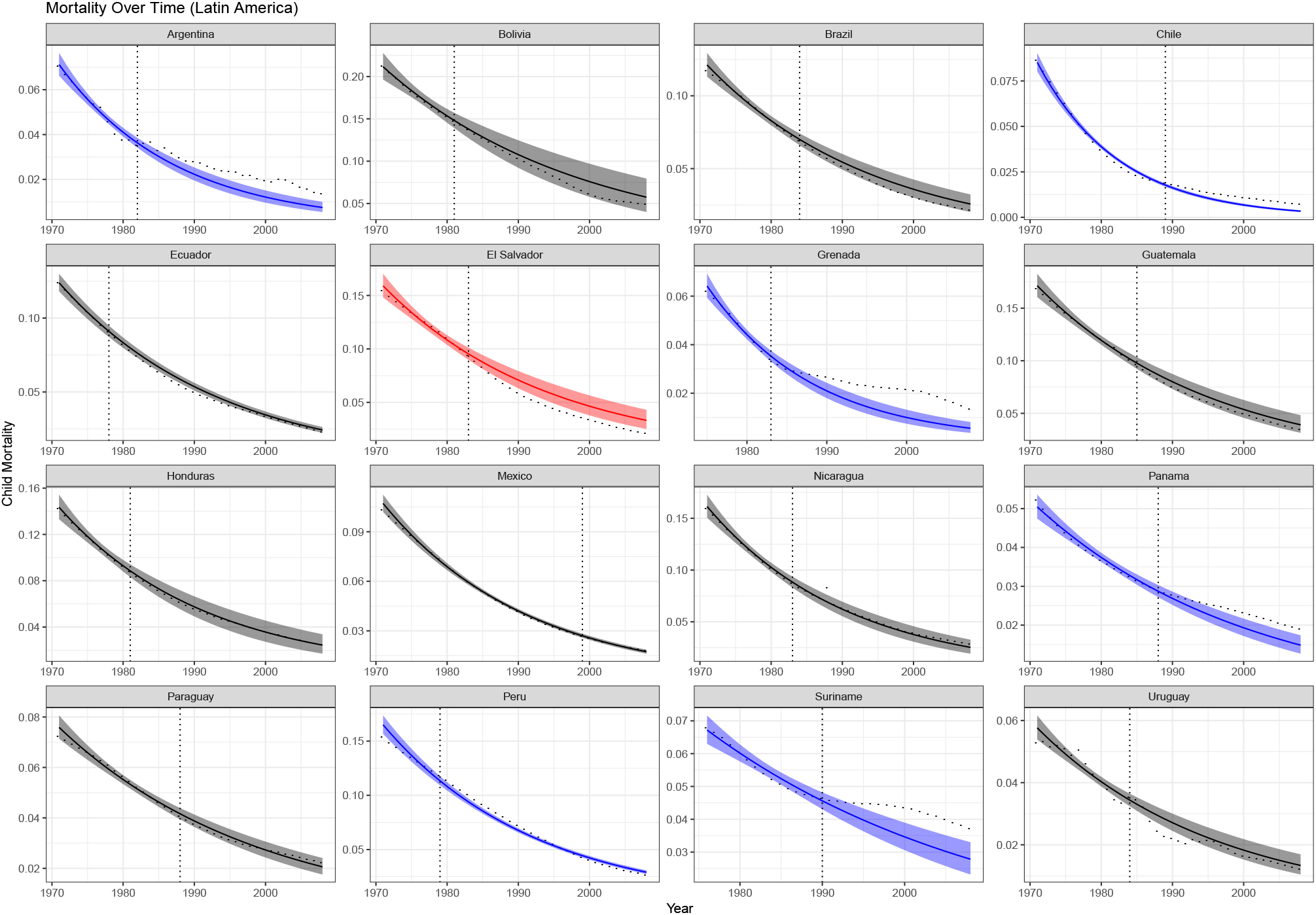
Empirical mortality rate (dots) and counterfactual mortality rates (solid line with shaded region) for Latin American countries. Beneficial effects are indicated by red counterfactual regions, harmful effects by blue, and neutral effects by grey. Dashed vertical line at the year of transition into democracy.

Finally, Figure 5 shows the remaining sixteen democratizers. All transitions were either beneficial or neutral: child mortality rates fell more quickly in five countries (Armenia, Cyprus, Mongolia, The Philippines, and Sri Lanka), and showed no measurable change in the other eleven.

**Figure 5:**
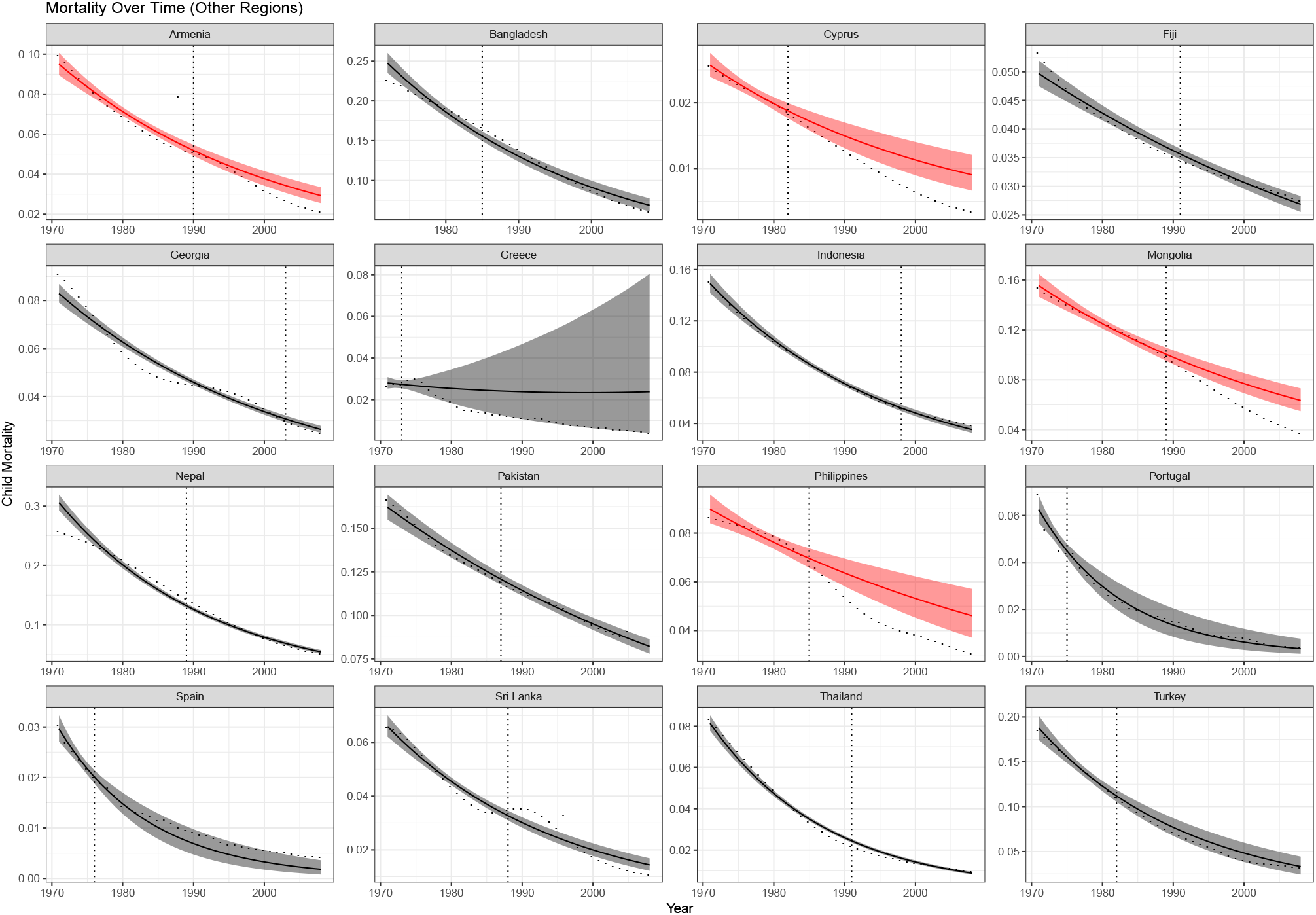
Empirical mortality rate (dots) and counterfactual mortality rates (solid line with shaded region) from other countries around the world. Beneficial effects are indicated by red counterfactual regions, harmful effects by blue, and neutral effects by grey. Dashed vertical line at the year of transition into democracy.

In addition to illustrate heterogeneous effects, these plots show that our model produces good in-sample fit. This is particularly useful for the pre-democratic period, as one of the important assumptions of Interrupted Time Series designs is a good in-sample fit for the pre-intervention period, which makes the extrapolation for the post transition period more credible (Morgan and Winship, 2019; Kontopantelis et al., 2015; Turner et al., 2019; Bernal et al., 2017).

### 3 Comparing Results for Child, Infant, and Neonatal Mortality

The models presented in the main text of the paper were all fit using the under-five (child) mortality rate as the outcome. Here we compare the results from child mortality with results obtained from models that use the infant (under 1 year) and neonatal (under 28 days) mortality rates as the outcomes. We look at short and long term effects.

First, we compare the estimates of the fixed effects and random effect variances between models that use child, infant, and neonatal mortality as the outcomes (Table 3). As expected, the intercept terms differ between the three models because, by definition, infants and neonates are a subset of all deaths that occurred among children under five years old. The short and long term effects are the same out to two decimal places. Similarly, aside from the random intercept variance, all other random effect variances are the same out to three decimal places. Overall we find that the results from the three models are very similar.

**Table 3:** Comparison of the effects of democracy on child mortality, infant mortality, and neonatal mortality as the outcomes. All results are based on the model from equation 1 in the paper.

| Parameter | Child | Infant | Neonatal |
| --- | --- | --- | --- |
| Intercept | -2.396 (-2.534, -2.257) | -2.68 (-2.794, -2.567) | -3.345 (-3.437, -3.256) |
| Time | -0.033 (-0.04, -0.027) | -0.032 (-0.038, -0.025) | -0.028 (-0.034, -0.022) |
| Short Term | 0.006 (-0.025, 0.036) | -0.001 (-0.035, 0.033) | -0.002 (-0.036, 0.03) |
| Long Term | -0.01 (-0.029, 0.01) | -0.01 (-0.029, 0.01) | -0.009 (-0.028, 0.01) |
| Random Effect Variances |  |  |  |
| Residual | 0.005 (0.005, 0.005) | 0.005 (0.004, 0.005) | 0.004 (0.003, 0.004) |
| Intercept | 0.854 (0.686, 1.058) | 0.584 (0.47, 0.731) | 0.374 (0.301, 0.468) |
| Time | 0.002 (0.001, 0.002) | 0.002 (0.001, 0.002) | 0.002 (0.001, 0.002) |
| Short Term | 0.013 (0.008, 0.019) | 0.014 (0.009, 0.021) | 0.013 (0.009, 0.02) |
| Long Term | 0.005 (0.004, 0.008) | 0.005 (0.004, 0.008) | 0.005 (0.004, 0.008) |

We now look at heterogeneity among in the effects of democracy for neonatal, infant, and child mortality. We compare the country-level intercepts and slopes in Figure 6. In each plot, the data have been sorted by the estimates from the model using child mortality as the outcome. We find that the intercept is highest in the child mortality models, followed by the infant mortality models and then the neonatal mortality models. In contrast, the ordering and magnitudes of the slopes are very similar for the three models. The ordering and magnitudes of the short term and long term effects are also very similar for the three models. Taken together, these results suggest that the results are similar for neonatal, infant, and child mortality.

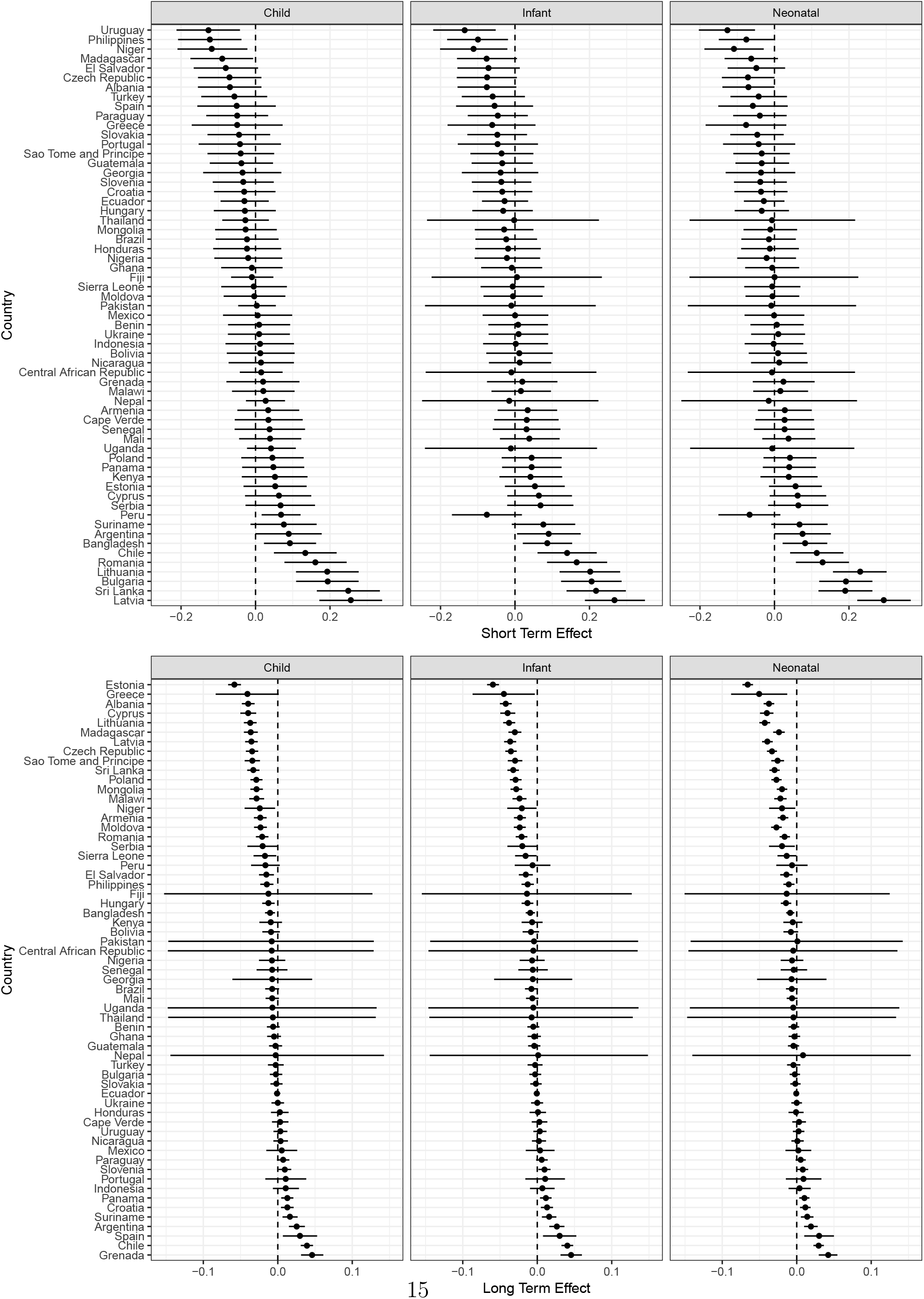
Figure 6

### 4 Foreign Aid in Sub-Saharan Africa

If foreign aid facilitates greater reductions in child mortality, it is due to increased pressure of resources from donors, and not for changes in the political system. Kudamatsu (2012) discussed how military coups against democratic governments in the 1990s were either suppressed by foreign intervention, such as in Comoros in 1995 and Lesotho in 1994 and 1998, or immediately followed by fresh multiparty elections due to donor pressure such as in Niger in 1996. Dunning (2004) found that it was only after the Soviet Union withdrew its financial support to African countries 1986 that the amount of Official Development Assistance (ODA) became positively correlated with the degree of democracy in Africa.

We use data on foreign aid from Kudamatsu (2012) to investigate whether the inclusion of foreign aid variables affects the results for Sub-Saharan Africa. The variables we include are total Official Development Assistant (ODA), ODA for the health sector (ODAH), and ODA for the water and sanitation (ODAWS). Figures 7, 8 and 9 display trends over time in foreign aid for each one of these variables. These figures show very little evidence that foreign aid flows increased after the introduction of democracy. In transitional countries where we do see a steady increase over time such as Malawi and Ghana, the post-democracy trend is very similar to the pre-democracy trend. Zambia and Mozambique also increase in ODA over time, although these countries were never democratic. Similar patterns can be found for ODAH and ODAWS.

**Figure 7:**
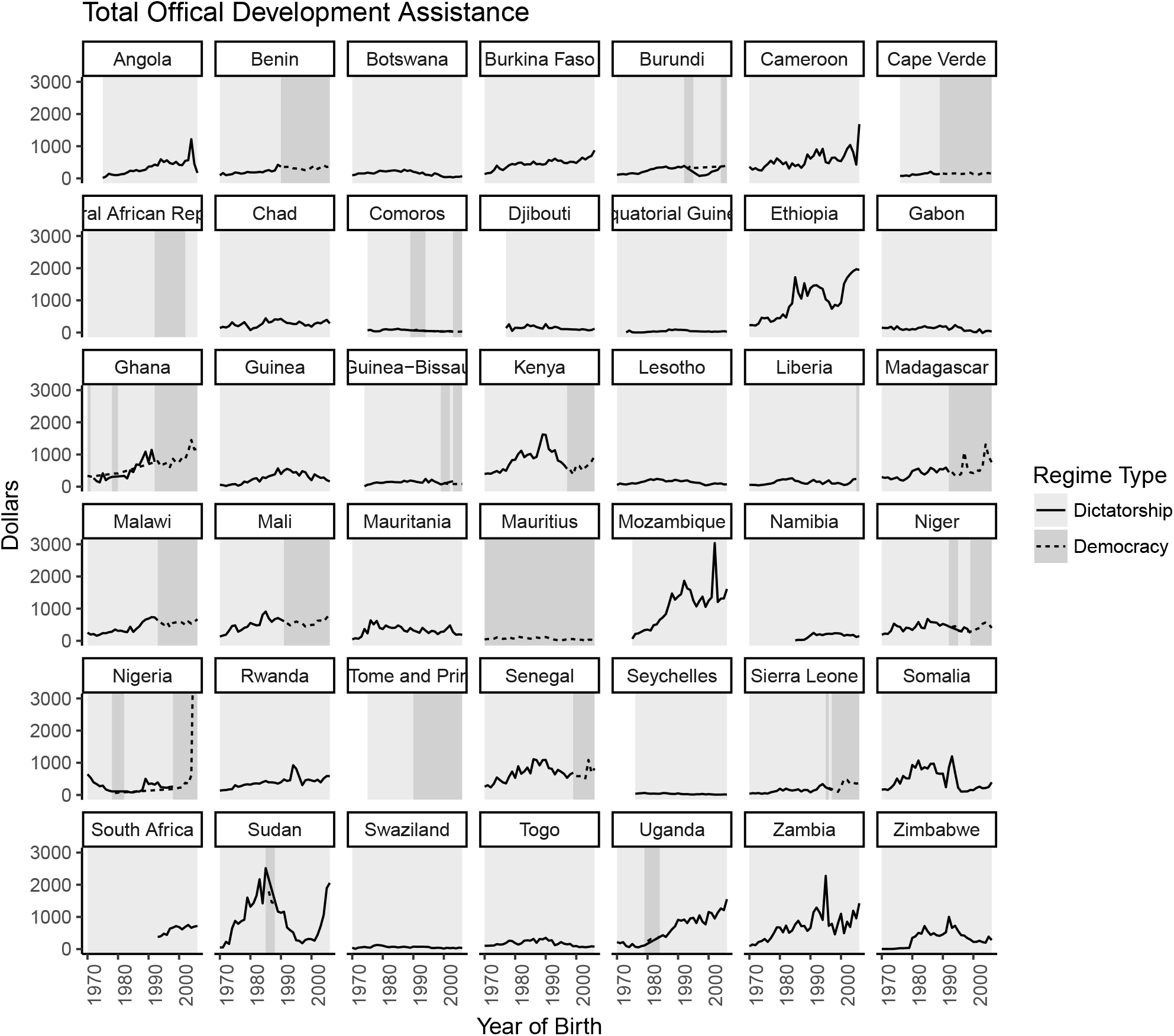
Total official development assistance by country in Sub-Saharan Africa.

**Figure 8:**
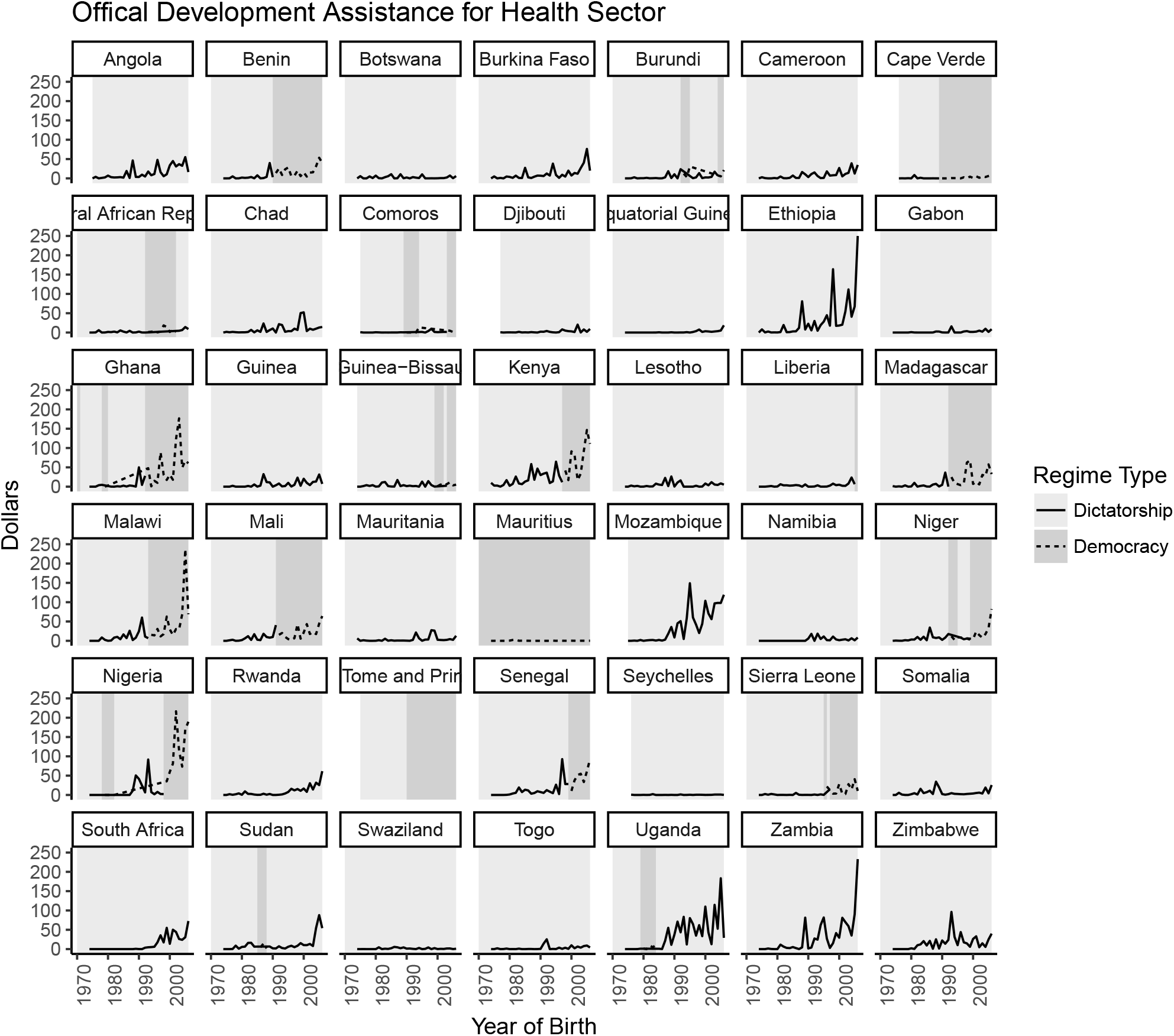
Official development assistance dedicated to the health sector in Sub-Saharan Africa.

**Figure 9:**
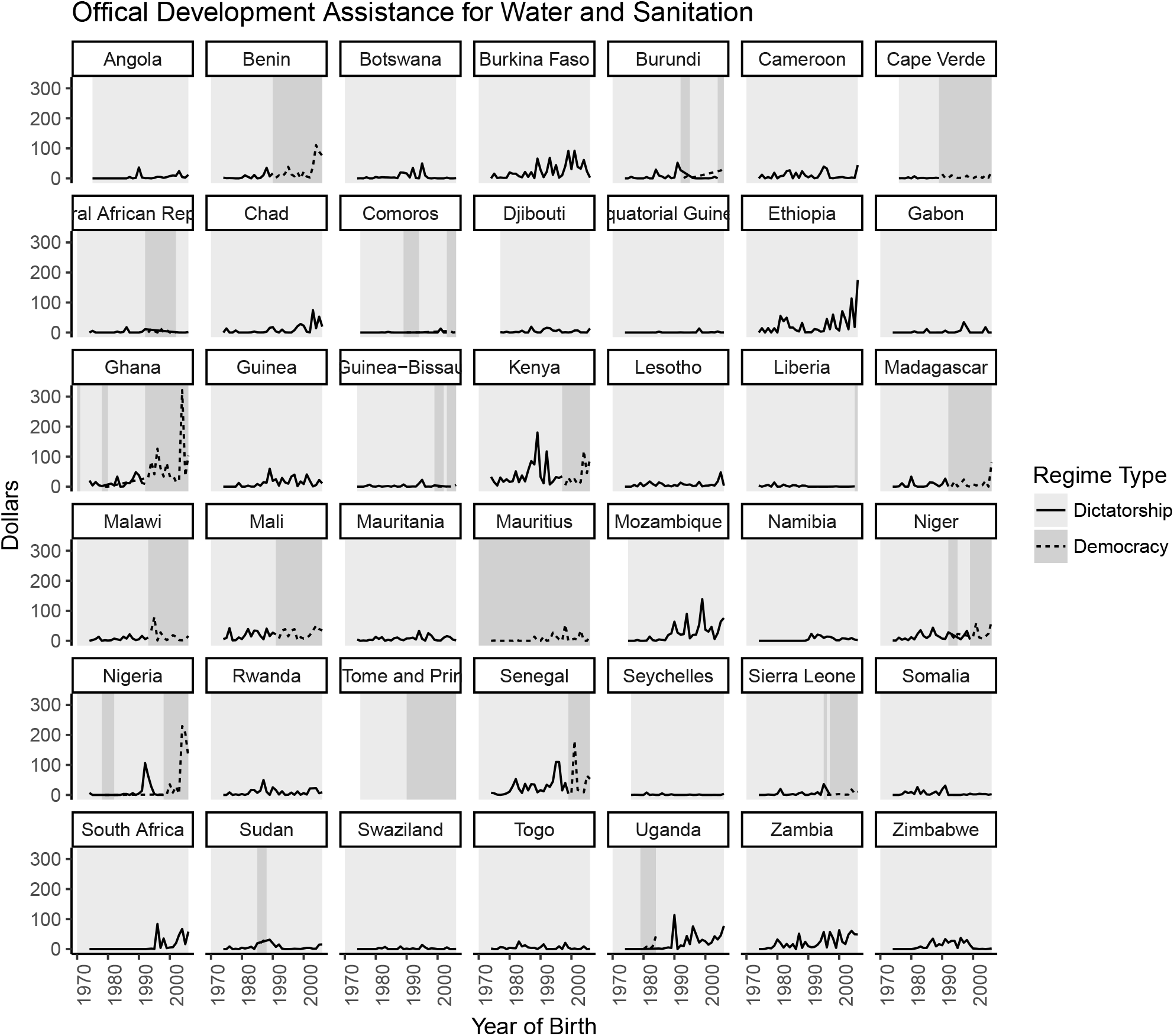
Offical development assistance dedicated to water and sanitation in Sub-Saharan Africa.

Table 4 shows the results for the regression models, where all foreign aid variables are lagged one year. We included country level random effects in all models, but these have been omitted from the table. Model 1 is our baseline model because it does not include any foreign aid variables. In Model 2, which includes total ODA, the short term effect seems to increase in magnitude, however, it also reduces the number of non-missing values from 1511 to 1400. To determine whether the effect on the short term effects is being driven by the ODA variable or reduced missing data, we fit Model 1 to the 1400 observations in Model 2. This model, which we call Model 3 shows that differences between Models 1 and 2 are largely due to the missing data. We repeat the same exercise using the additional variables ODAH (Models 4 and 5) and ODAWS (Models 6 and 7). In all cases the change in the regression coefficients are due to the missing data, not to the introduction of the variables related to foreign aid. Table 6 shows the details of the missing data.

**Table 4:** This table illustrates the effect of adding the predictors for foreign aid (ODA, ODAH,ODAWS), lagged 1 year, on the regression coefficients from our preferred specification. Model 1 is the baseline model. Models 2, 4, and 6 include ODA, ODAH, and ODAWS respectively. Models 3, 5, and 7 fit the the baseline model to the non-missing data in models 2, 4, and 6, respectively.

|  | Model 1 | Model 2 | Model 3 | Model 4 | Model 5 | Model 6 | Model 7 |
| --- | --- | --- | --- | --- | --- | --- | --- |
| (Intercept) | -1.0154***<br>(0.0948) | -1.0023***<br>(0.0963) | -0.9992***<br>(0.0962) | -1.0328***<br>(0.0966) | -1.0332***<br>(0.0966) | -1.0332***<br>(0.0967) | -1.0332***<br>(0.0966) |
| time | -0.0135***<br>(0.0020) | -0.0135***<br>(0.0020) | -0.0135***<br>(0.0020) | -0.0105***<br>(0.0021) | -0.0106***<br>(0.0021) | -0.0106***<br>(0.0021) | -0.0106***<br>(0.0021) |
| democracy.mjf | -0.0042<br>(0.0181) | -0.0049<br>(0.0200) | -0.0057<br>(0.0199) | -0.0052<br>(0.0181) | -0.0054<br>(0.0181) | -0.0054<br>(0.0181) | -0.0054<br>(0.0181) |
| new.time.mjf | -0.0085*<br>(0.0042) | -0.0055<br>(0.0042) | -0.0053<br>(0.0042) | -0.0059<br>(0.0040) | -0.0057<br>(0.0040) | -0.0057<br>(0.0040) | -0.0057<br>(0.0040) |
| maternal_educ | -0.0752***<br>(0.0159) | -0.0801***<br>(0.0162) | -0.0805***<br>(0.0163) | -0.1026***<br>(0.0167) | -0.1009***<br>(0.0166) | -0.1009***<br>(0.0166) | -0.1009***<br>(0.0166) |
| hiv_prev | 1.5964***<br>(0.0823) | 1.6631***<br>(0.0880) | 1.6647***<br>(0.0879) | 1.4700***<br>(0.0875) | 1.4744***<br>(0.0871) | 1.4743***<br>(0.0875) | 1.4744***<br>(0.0871) |
| log(IHME_usd_gdppc) | -0.0770***<br>(0.0115) | -0.0787***<br>(0.0119) | -0.0790***<br>(0.0119) | -0.0724***<br>(0.0119) | -0.0725***<br>(0.0119) | -0.0725***<br>(0.0119) | -0.0725***<br>(0.0119) |
| lag1.oda_real |  | 0.0000<br>(0.0000) |  |  |  |  |  |
| lag1.aid.health |  |  |  | 0.0001<br>(0.0001) |  |  |  |
| lag1.aid.water |  |  |  |  |  | 0.0000<br>(0.0001) |  |
| AIC | -3377.2372 | -3023.7579 | -3047.8734 | -2961.3705 | -2979.7230 | -2961.0156 | -2979.7230 |
| BIC | -3281.4677 | -2924.0905 | -2953.4516 | -2862.5341 | -2886.0885 | -2862.1792 | -2886.0885 |
| Log Likelihood | 1706.6186 | 1530.8790 | 1541.9367 | 1499.6852 | 1507.8615 | 1499.5078 | 1507.8615 |
| Num. obs. | 1511 | 1402 | 1402 | 1342 | 1342 | 1342 | 1342 |
| ***$p < 0.001$, **$p < 0.01$, *$p < 0.05$ | | | | | | | |

**Table 5:** Missing data for total official development assistance, development assistance for health and development assistant for health and sanitation.

| Country | ODAH | ODA | ODAW | ODAH | ODA | ODAW |  |
| --- | --- | --- | --- | --- | --- | --- | --- |
| Angola | 1 | 1 | 1 | Malawi | 5 | 0 | 5 |
| Benin | 5 | 0 | 5 | Mali | 5 | 0 | 5 |
| Botswana | 5 | 0 | 5 | Mauritania | 5 | 0 | 5 |
| Burkina Faso | 5 | 0 | 5 | Mauritius | 5 | 0 | 5 |
| Burundi | 5 | 0 | 5 | Mozambique | 0 | 0 | 0 |
| Cameroon | 5 | 0 | 5 | Namibia | 5 | 14 | 5 |
| Cape Verde | 0 | 0 | 0 | Niger | 5 | 0 | 5 |
| Central African Republic | 5 | 37 | 5 | Nigeria | 5 | 0 | 5 |
| Chad | 5 | 0 | 5 | Rwanda | 5 | 0 | 5 |
| Comoros | 0 | 0 | 0 | Sao Tome and Principe | 31 | 31 | 31 |
| Djibouti | 0 | 0 | 0 | Senegal | 5 | 0 | 5 |
| Equatorial Guinea | 5 | 4 | 5 | Seychelles | 0 | 0 | 0 |
| Ethiopia | 5 | 0 | 5 | Sierra Leone | 5 | 0 | 5 |
| Gabon | 5 | 0 | 5 | Somalia | 5 | 0 | 5 |
| Ghana | 5 | 0 | 5 | South Africa | 5 | 24 | 5 |
| Guinea | 5 | 0 | 5 | Sudan | 5 | 0 | 5 |
| Guinea-Bissau | 0 | 0 | 0 | Swaziland | 5 | 0 | 5 |
| Kenya | 5 | 0 | 5 | Togo | 5 | 0 | 5 |
| Lesotho | 5 | 0 | 5 | Uganda | 5 | 0 | 5 |
| Liberia | 5 | 0 | 5 | Zambia | 5 | 0 | 5 |
| Madagascar | 5 | 0 | 5 | Zimbabwe | 5 | 0 | 5 |

### 5 Autoregressive models and Random Effects for Time

Because of temporal correlation, log linear models for child mortality are often estimated by substituting independent normal errors with autoregressive process. Briefly, we say that ∊*_jt_* follows an autoregressive process of order *p* if,

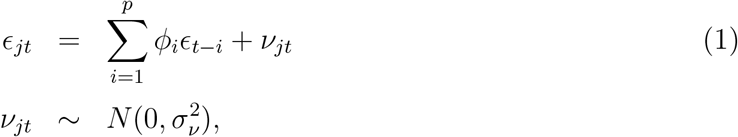

where appropriate constraints have been placed on the *φ_i_* to make the process stationary.

We experimented with this approach by modeling the residual errors in our model with an AR(1) process. However, this approach ended up not being feasible. For country *j*, let *Z_j_* be a design matrix with rows 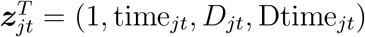 for *π_j_*. Also let *y_j_* = (*y_j_*_1_*,…, y_jnj_*) and ∊*_j_* = (*∊_j_*_1_*,…, ∊_jn_j__*), where *n_j_* is the number of time points for country *j*. Then the covariance matrix for *y_j_* is

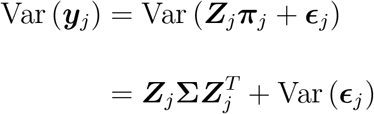

Thus, correlation between measurements in a country depends both on the random effects and the AR(1) correlation. These two components of the correlation are highly collinear, which makes the convergence of the model poor. This is a well-known problem in the spatial literature known as spatial confounding (Hodges and Reich, 2010). We experimented with different software, using the `lme` package in `R` for maximum likelihood estimation and JAGS and Stan for Bayesian estimation, and none of the models achieved convergence. Our code is available upon request.

Another issue is that the key quantities of interest in our analyses are the baseline mortality rates and trends over time for each country as well as the post-democracy deviations. Adding an auto-regressive structure to the models’ residual could potentially take a large part of the heterogeneity that we want to model via the random effects and treat it as unexplained correlation. Thus, even if it were possible to have both random effects and autoregressive processes in our model, it is not desirable to do so unless we believe that the random effects do not explain a large part of the within-country correlation.

## Footnotes

∗ Neonatal mortality is usually defined as deaths under 28 days of age; infant mortality as deaths under one year of age; and child mortality as deaths under 5 years of age.

† ? have updated these data to include more recent transitions in six additional countries, all in Africa: Burundi, Comoros, Lesotho, Liberia, Tunisia, and Zambia. Including these countries would usefully expand the transitions in our data by about ten percent and cast further light on the distribution of helpful, harmful, and neutral transitions in Africa. It would not change our basic finding: that the effects of democratic transitions are heterogeneous.

‡ A list of all the data sources is provided in the online appendix for Rajaratnam et al. (2010).

## References

Acemoglu, D., S. Johnson, J. Robinson, and P. Yared (2008). Income and Democracy. The American Economic Review 98 (3), 808–842.

Acemoglu, D., S. Naidu, P. Restrepo, and J. A. Robinson (2019). Does Democracy Cause Growth. Journal of Political Economy 127 (1), 47–100.

Aidt, T. S., J. Dutta, and E. Loukoianova (2006). Democracy comes to Europe: Franchise Extension and Fiscal Outcomes 1830–1938. European Economic Review 50 (2), 249–283.

Aidt, T. S. and P. S. Jensen (2009). The taxman tools up: An event history study of the introduction of the personal income tax. Journal of Public Economics 93 (1-2), 160–175.

Avelino, G., D. Brown, and W. Hunter (2004). Globalization and its Outcomes, Chapter Globalization, democracy, and social spending in Latin America, 1980-1997, pp. 209–228. Guilford Publications.

Banerjee, A., E. Duflo, R. Glennerster, and D. Kothari (2010). Improving immunisation coverage in rural India: clustered randomised controlled evaluation of immunisation campaigns with and without incentives. British Medical Journal 340 (may17 1), c2220.

Bassett, L. (2008). Can Conditional Cash Transfer Programs Play a Greater Role in Reducing Child Undernutrition? Social Protection Discussion Papers and Notes 46687, The World Bank.

Bell, A. and K. Jones (2015). Explaining Fixed Effects: Random Effects Modeling of Time-Series Cross-Sectional and Panel Data. Political Science Research and Methods 3 (1), 133–53.

Bernal, J. L., S. Cummins, and A. Gasparrini (2017). Interrupeted Time Series Regression for Evaluation Public Health Interventions: a tutorial. International Journal of Epidemiology, 348–55.

Besley, T. and M. Kudamatsu (2006a). Health and Democracy. American Economic Review 96 (2), 313–318.

Besley, T. and M. Kudamatsu (2006b). Health and Democracy. The American Economic Review 96 (2), 313–318.

Black, Robert, E., L. H. Allen, Z.q. A. Bhutta, L. E. Caulfield, M. d. Onis, M. Ezzati, C. Mathers, Rivera, Maternal, and C. U. S. Group (2008). Maternal and child undernutrition: global and regional exposures and health consequences. The Lancet 371, 243–60.

Black, R. E., S. S. Morris, and J. Bryce (2003). Where and Why are 10 million Children Dying Every Year? Commentary. Lancet 361, 2226–34.

Blaydes, L. and M. Kayser (2011). Counting Calories: Democracy and Distribution in the Developing World. International Studies Quartely 5, 887–908.

Bollyky, T. J., T. Templin, M. Cohen, J. L. Schoder, Diana Dieleman, and S. Wigley (2019, March). The relationships between democratic experience, adult health, and cause-specific mortality in 170 countries between 1980 and 2016: an observational analysis. The Lancet 393 10181 (10181), 1628–1640.

Brown, D. and W. Hunter (1999). Democracy and Social Spending in Latin America, 1980-92. American Political Science Review 93 (4), 779–790.

Bryce, J., S. e. Arifeen, G. Pariyo, C. F. Lanata, D. Gwatkin, J.-P. Habicht, and the Multi-Country Evaluation of IMCI Study Group (2003). Reducing Child Mortality: Can Public Health Deliver? The Lancet 362 (9378), 159–164.

Cheibub, J., J. Gandhi, and J. R. Vreeland (2010). Democracy and Dictatorship Revisited. Public Choice (2) (67–101.

de Kadt, D. d. and S. B. Wittels (2016). Democratization and Economic Output in Sub-Saharan Africa. Political Science Research and Methods 7 (1), 63–84.

De Mesquita, B. B., J. D. Morrow, and R. M. Siverson (2002). The Logic of Political Survival. British Journal of Political Science 32 (4), 559–590.

Devarajan, S. and R. Reinikka (2004). Making services work for poor people. Journal of African economies 13 (suppl 1), i142–i166.

Filmer, D. and L. Pritchett (1999). The Impact of Public Spending on Health: Does Money Matter? Social Science & Medicine 49 (10), 1309–1323.

Fitzmaurice, G., N. Laird, and J. Ware (2011). Applied Longitudinal Analysis. Wiley.

Flaxman, S., S. Mishra, A. Gandy, H. J. T. Unwin, T. A. Mellan, H. Coupland, C. Whittaker, H. Zhu, T. Berah, J. W. Eaton, M. Monod, and I. C. C.-. R. Team (2020). Estimating the effects of non-pharmaceutical interventions on COVID-19 in Europe. Nature.

Gelman, A. and J. Hill (2006). Data Analysis Using Regression and Multilevel/Hierarchical Model. New York: Cambridge University Press.

Gerring, J., S. C. Thacker, and R. Alfaro (2012a). Democracy and Human Development. Journal of Politics 74 (1), 1–17.

Gerring, J., S. C. Thacker, and R. Alfaro (2012b). Democracy and human development. The Journal of Politics 74 (1), 1–17.

Ghobarah, H. A., P. Huth, and B. Russett (2004). The post-war public health effects of civil conflict. Social science & medicine 59 (4), 869–884.

Gouveia, M. and N. A. Masia (1998). Does the median voter model explain the size of government?: Evidence from the states. Public Choice 97 (1-2), 159–177.

Gwatkin, D. R. (2004). Are Free Government Health Services The Best Way to Reach the Poor? Health, Nutrition and Population (HNP) Discussion Paper. http://siteresources.worldbank.org/HEALTHNUTRITIONANDPOPULATION/Resources/281627-1095698140167/Chapter2Final.pdf.

Harding, R. and D. Stasavage (2013). What Democracy Does (and Doesn’t Do) for Basic Services: School Fees, School Inputs, and African Elections. The Journal of Politics 76 (1), 229–245.

Houweling, T. A. and A. E. Kunst (2009). Socio-economic inequalities in childhood mortality in low- and middle-income countries: A review of the international evidence. British Medical Bulletin 93, 7–26.

Houweling, T. A., A. E. Kunst, C. W. Looman, and J. P. Mackenbach (2005). Determinants of under-5 mortality among the poor and the rich: a cross-national analysis of 43 developing countries. International journal of epidemiology 34 (6), 1257–1265.

Huber, E., T. Mustillo, and J. D. Stephens (2008). Politics and Social Spending in Latin America. The Journal of Politics 70 (2), 420–436.

Huber, E. and J. D. Stephens (2012). Democracy and the left: social policy and inequality in Latin America. University of Chicago Press.

Jones, G., R. W. Steketee, R. E. Black, Z. A. Bhutta, S. S. Morris, and the Bellagio Child Survival Study Group (2003). How Many Child Deaths Can We Prevent this Year? The Lancet 362 (9377), 65–71.

Justesen, M. (2012). Democracy, Dictatorship and Disease: Political Regimes and HIV/AIDS. Journal of European Political Economy 28 (3), 373–389.

Klasen, S. (2008). Poverty, undernutrition, and child mortality: Some inter-regional puzzles and their implications for research and policy. J Econ Inequal 6, 89–115.

Kohli, A. (2003). Democracy and development: Trends and prospects, Chapter Democracy, and Development. United Nations University Press.

Kontopantelis, E., T. Doran, I. Buchan, and D. Reeves (2015). CCBY Open access Research Methods Reporting Regression based quasi-experimental approach when randomisation is not an option: interrupted time series analysis. British Medical Journal 350.

Kosack, S. (2003). Effective Aid: How Democracy Allows Development Aid to Improve the Quality of Life. World Development 31 (1), 1–22.

Krueger, P. M., K. Dovel, and K. Denney (2015). Democracy and self-rated health across 67 countries: A multilevel analysis. Social Science & Medicine 143, 137–144.

Kudamatsu, M. (2012). Has Democratization Reduced Infant Mortality in Sub-Saharan Africa? Evidence from Micro Data. Journal of the European Economic Association 10 (6), 1294–1317.

Lake, D. A. and M. A. Baum (2001). The Invisible Hand of Democracy: Political Control and The Provision of Public Services. Comparative Political Studies 34 (6), 587–621.

Lindert, P. H. (2004). Growing public: Social spending and economic growth since the eighteenth century (2 ed.). Cambridge University Press.

Lipset, S. M. (1959). American intellectuals: their politics and status. Daedalus 88 (3), 460–486.

McGuire, M. C. and M. Olson (1996). The economics of autocracy and majority rule: the invisible hand and the use of force. Journal of economic literature 34 (1), 72–96.

McGuire, S. (2013). Multinationals and NGOs amid a changing balance of power. International Affairs 89 (3), 695–710.

Meltzer and Richard (1981). A Rational Theory of the Size of Government. Journal of Political Economy 89 (5), 814–927.

Min, B. (2015). Power and the Vote: Elections and Electricity in the Developing World. New Jersey, NY, USA: Cambridge University Press.

Moore, M. and H. White (2003). States, markets, and just growth: development in the twenty-first century, Chapter Meeting the challenge of poverty and inequality, pp. 64–95. 1: United Nations University Press.

Morgan, S. and C. Winship (2019). Counterfactual and Causal Inference: Methods and Principles for Social Science Research (second ed.). Cambridge University Press.

Mulligan, C. B., R. Gil, and X. Sala-i Martin (2004). Do Democracies Have Different Public Policies than Nondemocracies? Journal of Economic Perspectives 19 (1), 51–74.

Niskanen, W. A. (1997). Autocratic, democratic, and optimal government. Economic Inquiry 35 (3), 464–479.

Olper, A., J. Falkowki, and J. Swinnen (2013). Poltical Reforms and Public Policy: Evidence from Agricultural and Food Policies. The World Bank Economic Review 28 (1), 21–47.

Pateman, C. (1976). Participation and Democratic Theory. Cambridge University Press.

Persson, T., G. E. Tabellini, et al. (2003). Do electoral cycles differ across political systems? Innocenzo Gasparini Institute for Economic Research.

Peyvand, G. and V. Gauri (2002). Immunization in developing countries: its political and organizational determinants. World Development 30 (12), 2109–2132.

Pieters, H., D. Curzi, A. Olper, and Swinnen (2016). Effect of democratic reforms on child mortality: a synthetic control analysis. The Lancet: Global Health 4 (9), 627–632.

Pritchard, C. and S. Keen (2016). Child mortality and poverty in three world regions (the West, Asia and Sub-Saharan Africa) 1988–2010: Evidence of relative intra-regional neglect? Child mortality and poverty in three world regions (the West, Asia and Sub-Saharan Africa) 1988–2010: Evidence of relative intra-regional neglect? Scandinavian Journal of Public Health 44, 734–741.

Przeworski, A., M. Álvarez, J. Cheibub, and F. Limongi (2000). Democracy and Development: Political Institutions and Well-Being in the World, 1950–1990. New York: Cambridge University Press.

Putman, R. (1993). Making Democracy Work: Civic Traditions in Modern Italy. Princeton University Press.

Rajaratnam, J. K., J. R. Marcus, A. D. Flaxman, H. Wang, A. Levin-Rector, L. Dwyer, M. Costa, A. D. Lopez, and C. J. L. Murray (2010). Neonatal, Postneonatal, Childhood, and Under-5 Mortality for 187 countries, 1970-2010: a Systematic Analysis of Progress Towards Millennium Development Goal 4. The Lancet 375 (9730), 1988–2008.

Rasella, D., Basu, T. Hone, R. Paes-Sousa, C. O. Ocke-Reis, and C. Millett (2019). Child morbidity and mortality associated with alternative policy responses to the economic crisis in Brazil: A nationwide microsimulation study. PLoS Medicine 5, e1002570.

Reinsberg, B. (2015). Foreign aid responses to political liberalization. World Development 75, 46–61.

Reisel, G. (1985). Mean Squared Error Properties of Empirical Bayes Estimators in a Multivariate Random Effects General Linear Model. Journal of American Statistical Association 80, 642–650.

Robinson, G. K. (1991). That BLUP is a Good Thing: The Estimation of Random Effects. Statistical Science 6 (1), 15–51.

Ross, M. (2006). Is Democracy Good for the Poor? American Journal of Political Science 50 (4), 860–874.

Safaei, J. (2012). Pos-Comunist Health Transitions in Central and Eastern Europe. Economics Research International.

Sen, A. (1999a). Development as Freedom. Oxford University Press paperback. OUP Oxford.

Sen, A. K. (1999b). Democracy as a universal value. Journal of democracy 10 (3), 3–17.

Shandra, J., J. Nobles, and B. London (2004). Dependency, democracy, and infant mortality: a quantitative, cross-national analysis of less developed countries. Social Science & Medicine 59, 321–333.

Singer, J. D. and J. B. Willett (2003). Applied Longitudinal Data Analysis: Modeling Change and Event Occurrence. Oxford University Press.

Stasavage, D. (2005). Democracy and Education Spending in Africa. American Journal of Political Science 49 (2), 343–358.

Stigler, G. J. (1970). Director’s Law of Public Income Redistribution. Journal of Law and Economics 13 (1), 1–10.

Svensson, J. (1999). Aid, Growth and Democracy. Economics and Politics 11, 275–297.

Tavares, J. and R. Wacziarg (2001). How democracy affects growth. European economic review 45 (8), 1341–1378.

Turner, S. L., A. Karahalios, A. B. Forbers, M. Tajaard, J. M. Grimshaw, A. C. Cheng, L. Bero, and J. McKenzie (2019). Design characteristics and statistical methods used in interrupted time series studies evaluation public health interventions: protocol for review. British Medical Journal Open 9.

unicef (1997). Chidren at Risk in Central and Eastern Europe: Perils and Promisses. Reginal Monitoring Report. https://www.unicef-irc.org/publications/48-children-at-risk-in-central-and-eastern-europe-perils-and-promises.html.

Victora, C. G., A. Wagstaff, Armstrong, J. Schellenberg, D. Gwatkin, M. Claeson, and J.-P. Habicht (2003). Applying an Equity Lens to Child Health and Mortality: More of the Same is Not Enough. The Lancet 362 (9379), 233–241.

Vokó, Z. and J. G. Pitter (2020). The effect of social distance measures on COVID-19 epidemics in Europe: an interrupted time series analysis. GeroScience 42, 1075–1082.

Wang, Y., V. Mechkova, and F. Andersson (2018). Does Democracy Enhance Health? New Empirical Evidence 1900–2012. Political Research Quarterly 00 (0), 1–16.

Weiss, R. E. (2005). Modeling longitudinal data. Springer.

Wigley, S. and A. Akkoyunlu-Wigley (2011). The impact of regime type on health: does redistribution explain everything? World Politics 63 (4), 647–677.

Wigley, S. and A. Akkoyunlu-Wigley (2017). The impact of democracy and media freedom on under-5 mortality, 1961e2011. Social Science & Medicine 190, 237–246.

## References

Cheibub, J., J. Gandhi, and J. R. Vreeland (2010). Democracy and Dictatorship Revisited. Public Choice (67–101.

Dunning, T. (2004, April). Conditioning the Effects of Aid: Cold War Politics, Donor Credibility, and Democracy in Africa. International Organization 58, 409–423.

Hodges, J. and B. Reich (2010). Adding Spatially-Correlated Errors Can Mess Up the Fixed Effect You Love. Journal of American Statistical Association 64 (4), 325–334.

